# 1Hz vs. cTBS Transcranial Magnetic Stimulation to Augment Behavior Therapy for Tics: A Randomized Clinical Trial

**DOI:** 10.64898/2026.09.29.26364036

**Authors:** Christine A. Conelea, Sarah Kretsch, Deanna J. Greene, Bryon A. Mueller, Claire Breitenfeldt, Kerry Houlihan, Brianna Wellen, Timothy Hendrickson, Jennifer Alexander, Mark Fiecas, Erjia Cui, Kelly Duffy, David Schneck, Mo Chen, Charles P. Lewis, Donovan Roediger, Alexander Opitz, Suma Jacob, Kelvin Lim

**Author notes:** **Corresponding Author**: Christine Conelea. CB is now affiliated with the University of Utah, Department of Psychology; KH is now affiliated with Bowling Green State University, Department of Psychology; JA is now affiliated with Psychology Houston; MC is now affiliated with Gillette Children’s Hospital; SJ is now affiliated with the University of California, Los Angeles, Semel Institute, Department of Psychiatry & Biobehavioral Sciences.

## Abstract

**Background:** Response to Comprehensive Behavioral Intervention for Tics (CBIT), the first-line treatment for tic disorders, is suboptimal. We conducted Phase 1 of a two-phase, milestone-driven trial to determine if augmenting CBIT with repetitive transcranial magnetic stimulation (TMS) to supplementary motor area (SMA) could safely engage targeted sensorimotor circuitry and improve tic controllability.

**Methods:** In a 3-arm, double-blind, sham-controlled randomized trial (NCT04578912), 50 youth (ages 12–21) with moderate-to-severe tics received 10 sessions of either 1Hz rTMS, continuous theta burst stimulation (cTBS), or sham stimulation to the SMA, immediately followed by CBIT. Primary outcomes were safety, feasibility, and neural target engagement (fMRI SMA activation and resting-state functional connectivity [RSFC]). Secondary outcomes included change in tic controllability, tic frequency at rest, and urges.

**Results:** The CBIT+TMS intervention was safe and feasible (96% completion). Treatment-related adverse events were mild-to-moderate and occurred more frequently in the 1Hz rTMS group (27% of sessions) vs. cTBS (14.7%) and sham (11.3%). While task-based SMA activation remained unchanged, cTBS demonstrated robust neural target engagement, reducing SMA-primary motor cortex (M1) RSFC and putamen-M1 RSFC. Conversely, 1Hz rTMS blunted the cortico-striatal connectivity reductions observed in the sham group. All groups exhibited significant reductions in tic frequency and improved controllability. Premonitory urges increased in the 1Hz rTMS group but decreased in sham and cTBS.

**Conclusions:** SMA-directed cTBS is a safe, feasible augmentation that may amplify the neural effects of CBIT on sensorimotor circuitry. Findings identify cTBS as the preferred stimulation regimen for future efficacy testing.

**Clinical Trial Registration:** ClinicalTrials.gov; Identifier: NCT04578912.

## INTRODUCTION

Tourette Syndrome (TS) and chronic tic disorders are child-onset neurodevelopmental conditions affecting 1-3% of people (1). Tics are unwanted, repetitive movements and vocalizations that can be preceded by premonitory urges and suppressed for short periods of time. Tics can adversely affect functioning and interfere with daily tasks (2,3).

Large randomized control trials (4,5) and meta-analyses (6) support Comprehensive Behavioral Intervention for Tics (CBIT; (7)) as the first-line treatment for tics (8). CBIT focuses on enhancing a patient’s ability to engage voluntary control over functionally impairing tics (“tic controllability”). However, many youth lack this foundational ability (9) and only ∼50% of children (4) and ∼40% of adults benefit (5). Augmentation targeting neurocircuitry involved in tic control may boost CBIT response.

Tics are proposed to arise from dysfunctional striatal activity that generates widespread effects across cortico-striatal-thalamo-cortical circuitry (CSTC)(10,11), including in the supplementary motor area (SMA). SMA plays a central role in motor inhibition, including proactive inhibitory control of movement initiation and reactive control/movement cancellation (12). In TS, SMA shows increased activation prior to tics (10,13,14) and during tic periods (14,15), and it has enhanced functional connectivity with primary motor cortex (M1),(14),(16) and the putamen (17). These abnormalities correlate with clinical ratings of premonitory urge intensity (18) and tic severity (19). In people without tics, SMA stimulation can induce tic-like movements and urges (20,21).

SMA has emerged as a leading candidate target for tic intervention using repetitive transcranial magnetic stimulation (TMS) “inhibitory” paradigms (22). While some open-label studies of 1Hz rTMS to SMA (23–25) reported reduced clinician-rated tic severity on the Yale Global Tic Severity Scale (YGTSS; (26)), randomized trials targeting SMA with continuous theta burst stimulation (cTBS)(27) and deep TMS (HBDL coil; (28)) did not find such change. Varied effects may reflect methodological differences in TMS paradigms and dosing. For example, a trial with null effects after 3 weeks of daily 1Hz rTMS to SMA found significant tic reduction after 3 additional weeks of treatment (29).

To date, no TMS trials for tics have directly compared stimulation paradigms, examined effects on tic controllability, or considered the brain-state dependency of TMS effects (30). Neural mechanisms also remain underexplored. In the only trial that included fMRI, Wu et al. (27) observed significant decreases in SMA activation and connectivity to bilateral M1 after 2 days of cTBS, suggesting stimulation may modulate sensorimotor circuitry “toward health.”

To address these gaps, we designed the “CBIT+TMS trial,” a two-phase, milestone-driven, early-stage randomized controlled trial to test whether augmenting CBIT with 1Hz rTMS or cTBS to the SMA reduces SMA activation and resting state functional connectivity (RSFC) and enhances tic controllability compared to sham in youth with tics (31). Here we report Phase 1 primary outcomes, which tested: 1) TMS safety and feasibility, 2) neural target engagement and 3) behavioral target engagement. We hypothesized that 1) TMS would be safe and feasible, with a treatment completion rate of ≥80% and ≤20% rate of treatment-related adverse events; 2) either or both active TMS groups would show more pronounced change than sham on at least one fMRI-measured neural target (medium to large within-subject effect size of ≥ η2=0.18 for change in SMA motor task activation *or* RSFC within sensorimotor circuit nodes); and 3) either or both active TMS groups would show greater improvement in tic controllability vs. sham.

## METHODS AND MATERIALS

### Design

This 3-arm, randomized, double-blind, sham-controlled trial was conducted in an outpatient clinical research setting between November 2020-December 2023. The study was preregistered (Clinicaltrials.gov: NCT04578912), and the protocol and a priori hypotheses were published prior to completion of data collection (31). The study was approved by the University of Minnesota Institutional Review Board, and an Investigational Device Exemption was granted by the U.S. Food and Drug Administration. Prior to data collection, adults provided written consent and minors written assent plus written consent from a parent/guardian. Youth who turned 18 years old during their participation were re-consented as adults.

### Participants

Eligibility was confirmed following an assessment and medical record review. <u>Inclusion criteria</u> were: 1) age 12-21 years (inclusive), 2) current tics of ≥1 year duration (32) and 3) at least moderate tic severity (YGTSS (26) total score ≥14 or ≥9 for those with motor or vocal tics only), 4) IQ >70 on the Wechsler Abbreviated Scale of Intelligence (WASI; (33)), and 5) English fluency. <u>Exclusion criteria</u> were: 1) conditions contraindicated or associated with increased TMS risk profile (e.g., intracranial pathology, seizure disorders, implanted medical devices, pregnancy), 2) inability to undergo MRI, 3) left handedness, 4) active suicidality, 5) previous diagnosis of psychosis or cognitive disability, 6) substance abuse or dependence within the past year, 7) concurrent psychotherapy focused on tics, and 8) current neuroleptic medication. Other psychotropic medications were allowed if stable ≥6 weeks with no anticipated changes.

### Procedures

#### Assessments

Assessment visits occurred at pre-treatment (within 10 calendar days before the first treatment session), post-treatment (within 10 calendar days after final treatment session), and 1- and 3-month follow-ups. Clinical measures were administered to inform eligibility, characterize the sample, and assess for possible iatrogenic effects. Assessments included self-/parent-report questionnaires and clinician-report measures administered by an independent evaluator (IE) masked to TMS condition and CBIT progress. Measures included the YGTSS (26), Premonitory Urge for Tics Scale (PUTS; (34)), MINI-KID 7 (35) or MINI 7 (36), Clinical-Global Impressions-Improvement (CGI-I ((37)), TMS Adverse Effects Questionnaire (38), and Client Satisfaction Questionnaire-8 (39) (see Supplement for details and relevant results).

The Tic Suppression Task (TST) (9,40,41) is an established direct observation paradigm that was used at each assessment timepoint to measure tic frequency (tics per minute; behavioral target) and mean urge ratings. A video (42) instructed participants during two 3-minute conditions of 1) “Free-to-Tic” (naturally occurring tic frequency) and 2) “Suppression” (attempted tic inhibition). A visual 0-10 scale appeared every 30 seconds to prompt a verbal rating of premonitory urge intensity. TSTs were video recorded and coded by trained raters masked to TMS condition, TST condition instructions, and study time point. Inter-rater reliability was established (Supplement).

#### MRI Data

MRI data were acquired on a Siemens Prisma 3T scanner with 32-channel receive-only head coil at pre- and post-treatment. Post-treatment scans occurred at least 24 hours after the last TMS session. The Adolescent Brain Cognitive Development (ABCD) study acquisition protocol (43) was used for structural (T1, T2), resting-state fMRI, and task fMRI (bilateral finger tapping during 15 second blocks of tap vs. rest, to functionally localize SMA) data collection (parameters in Supplement).

#### Interventions

Participants received 10 treatment sessions delivered daily on weekdays for 2 weeks. Each treatment visit consisted of TMS immediately followed by the CBIT session.

#### TMS

TMS was delivered with a Magstim Super Rapid2 stimulator with a Magstim air-cooled 70mm figure-eight coil for resting motor threshold (RMT) determination and active conditions and a Magstim sham air-cooled coil for the sham condition. RMT was measured immediately before the first treatment session using single-pulse TMS to the left M1 hand area. RMT was defined as the minimum magnetic flux eliciting a threshold EMG response (≥ 50 μV in peak-to-peak amplitude) in a resting target muscle (abductor pollicis brevis) in 5/10 trials. Treatment parameters were: 1) 1 Hz: single train of 2000 pulses, 110% RMT (33 minute duration); 2) cTBS: bursts of 3 pulses at 30 Hz repeated every 200 ms (5 Hz burst frequency), single uninterrupted 40 second train, 600 total pulses, 90% RMT (40 second duration); (3) sham: audible stimuli only mimicking active TMS; half of participants received sham mimicking the 1 Hz sequence and half the cTBS sequence. TMS targets were computationally defined using individual-level structural MRI, task fMRI, and finite electric field modeling data (44; Supplement).

#### CBIT

CBIT followed the published manual (7) and was delivered by therapists trained and supervised by a licensed psychologist. CBIT components are: 1) psychoeducation; 2) functional interventions; 3) competing response training (using a competing motor action contingent on tics or tic antecedents, taught in a graded manner, beginning with the most distressing tics); and 4) social support. The protocol emphasized therapist-supported practice of competing responses. Competing responses practice was assigned for homework. Fidelity ratings indicated strong adherence (Supplement).

### Randomization

Block randomization was stratified on TST performance (change in tic frequency from Free-to-Tic to Suppress conditions below or above 50%) and medication status (on vs. off) using the R blockrand package (45). Blocks were sizes of 6, designed so that odds of allocation were 1:1 for 1Hz vs. cTBS and 2:1 for active vs. sham.

### Blinding

TMS status (active vs. sham) was only known to the statistician and TMS operator supervisor; participants, investigators, and all other study staff were masked. TMS operators, participants, and parents were aware of 1 Hz vs. cTBS allocation due to different session duration. Active vs. sham coils were concealed with coded letters, and a separate active coil was used for RMT. Integrity of the blindwas adequate (see Supplement).

### Analysis Plan

Primary outcomes were: 1) safety and feasibility, indexed by rates of treatment completion and TMS-related adverse events and 2) neural change, defined as within-subject change from pre- to post-treatment in participants with adequate quality imaging data in a) SMA activation, as assessed by BOLD signal during an fMRI motor task (bilateral finger tapping) and b) RSFC of SMA-mediated brain circuits (SMA-Putamen, SMA-M1). Secondary outcomes measured through 3-month follow-up were TST changes in tic frequency, tic controllability, and urges. Analysis plans were pre-registered and previously described fully (31); approach is summarized here.

#### Safety and feasibility

Rates of AEs judged “likely” or “possibly” TMS-related were calculated at the 1) person-level (N of participants who experienced the AE at least once / total N of participants in the arm) and 2) session-level (N of sessions with AEs reported / total N of treatment sessions). Descriptive statistics were used to examine feasibility (treatment completion rates) and intervention acceptability (Client Satisfaction Questionnaire-8; CSQ-8; (39)).

#### Neural target engagement

##### SMA activation

BOLD activation parameters in an SMA ROI (5 mm radius sphere) were identified for each participant in a first-level general linear model analysis for the contrast tap vs. rest. Activation parameters were entered into a linear mixed effects model (LMM) to examine whether within-subject change in SMA activation on the fMRI motor task from pre- to post-treatment differed by group.

##### SMA connectivity

A seed-based analysis approach tested RSFC between a priori ROIs (SMA, putamen, M1 hand and mouth areas, representing the largest M1 regions) using a false discovery rate correction for multiple comparisons threshold of *p*<0.05. ROIs were individually anatomically defined using each participant’s T1 and the Steitzman et al. (46) parcellation scheme aligned to their RSFC. As multiple atlas parcels fall within SMA region, the parcel containing the individual’s TMS target was used for the SMA ROI in connectivity analyses (see Fig. 1). Left hemisphere ROIs for M1 areas and putamen were selected in the connectivity analysis with the SMA ROI because TMS stimulation intensity was based on left motor cortex and predominance of left hemisphere motor areas was presumed given participant right handedness.

For each participant at each timepoint, average BOLD time-series data were extracted from each ROI and used to calculate correlation coefficients between ROIs. These were converted into individual z-scores using Fischer’s transformation. ANOVA was used to model within-subject change in RSFC from pre- to post-treatment by TMS group. An ANOVA effect size of ≥ χ2 = 0.18 was set a priori as the target engagement threshold. Cohen’s *d* was calculated to understand the direction of the effect (i.e., increased or decreased functional connectivity).

**Figure 1.**
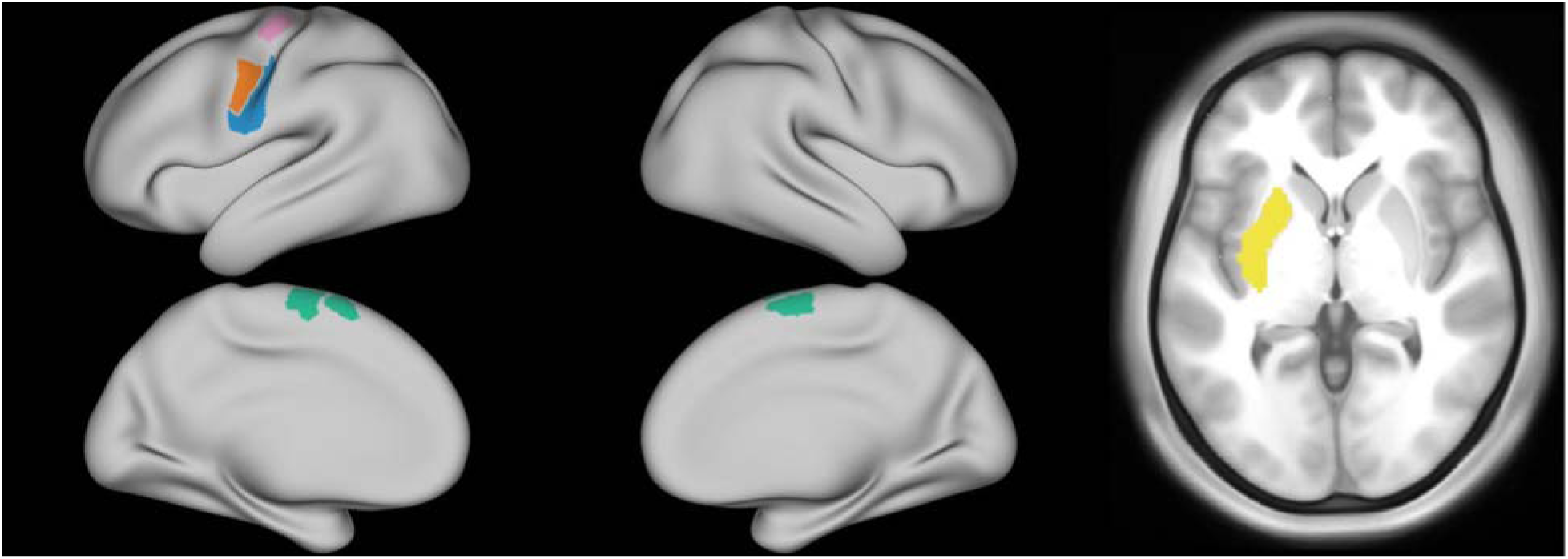
Regions of interest for resting state functional connectivity analyses. Yellow = putamen. Green = Three of four SMA parcels in which TMS targets fell (parcel 4 is hidden on the right superior cortex); for analyses, the SMA ROI used was the parcel in which an individual’s TMS target was located. Pink = L M1 Hand 10; Orange = M1 Mouth 1; Blue = M1 Mouth 3.

#### Behavioral target engagement

LMMs characterized changes in mean TST tic frequency across timepoints. Two separate models were fit for each condition (Free-to-Tic, Suppression) to capture acute (pre, post) and long-term results (1- and 3-month follow-ups). Main predictors of interest were group effect and group by time interaction effects. TST Suppression models included a term for mean pre-treatment tic frequency in the TST Free-to-Tic condition. Follow-up models included a term for pre-post mean tic change. Each LMM included a random intercept term for participant ID to account for within-subject correlation between visits. All mean tic outcomes were deemed non-normal, so a logarithm transformation was implemented to reduce skewness.

The same LMM procedure was used for mean urges across timepoints for TST conditions. Outcome was defined as mean urges across each session. Main predictors of interest were the group effect and the group by time effect.

## RESULTS

Recruitment and participant flow are illustrated in the CONSORT diagram (Supplement). The randomized sample consisted of 50 youth ages 12-21 years (M = 16.01 years, SD = 1.14), with n = 16 assigned to cTBS, n = 17 to 1 Hz rTMS, and n = 17 to sham. Baseline demographic and clinical characteristics are in Table 1.

**Table 1.**
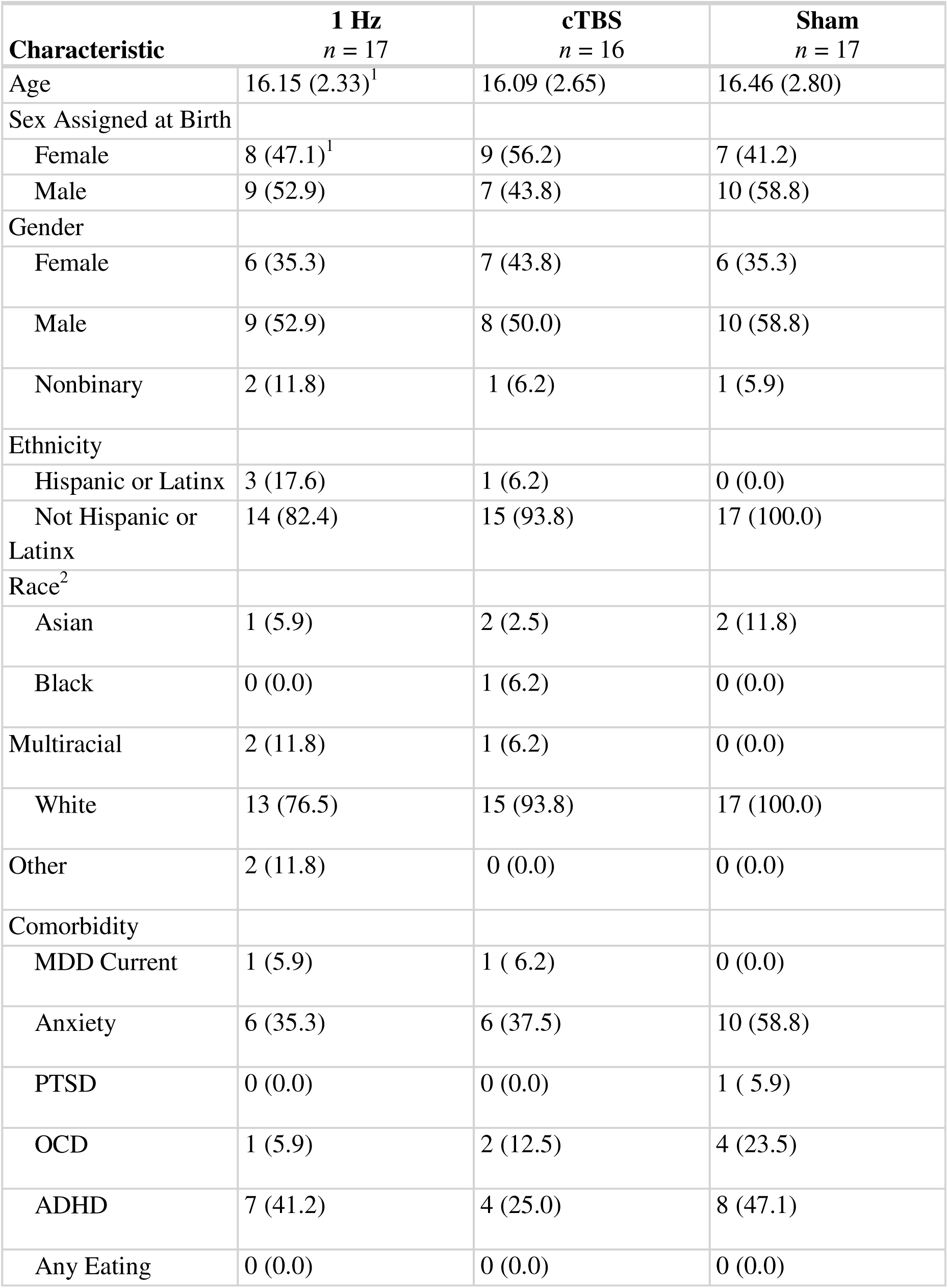
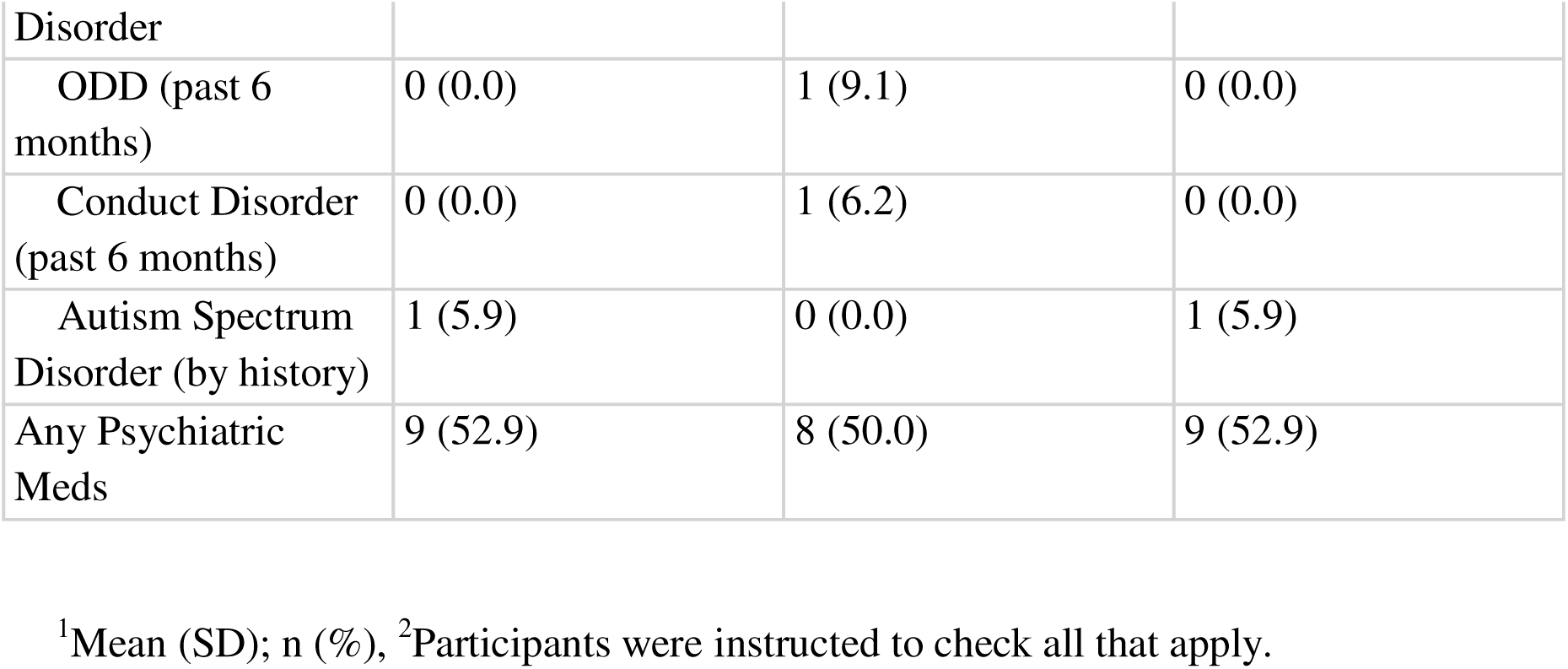
Demographic and clinical characteristics of randomized participants, by group and total.

### Safety and feasibility

Treatment was completed by 48 (96%) participants. None withdrew due to TMS tolerability issues. All completers were retained through 3-month follow-up.

There were no serious AEs and no AEs rated as “severe” and TMS-related (i.e., all treatment-related AEs were “mild” or “moderate”). Specific TMS-related AE rates are reported in the Supplement. The number of participants who experienced at least one TMS-related AE was similar in both active conditions (cTBS: n = 11, 73.3%; 1 Hz: n = 12, 70.6%) and higher than sham (n = 8, 50%). The number of sessions with a TMS-related AE was higher in 1 Hz (n = 46, 27%) than in both sham (n = 18, 11.3%) and cTBS (n = 22, 14.7%). Rates of specific AEs were similar to those reported previously (47), and none exceeded a rate of 20%.

### Intervention acceptability

Treatment satisfaction was rated highly and similarly across groups on the CSQ-8 self-report (1Hz: M = 28.76, SD = 3.75; cTBS: M = 28.93, SD = 2.34; sham: M = 29.06, SD = 2.65) and parent-report (*n* = 36; 1Hz: M = 30.15, SD = 2.82; cTBS: M = 29.09, SD = 3.18; sham: M = 29.92, SD = 2.81).

### Neural target engagement. SMA activation

Analyses did not confirm the hypothesis that either or both active TMS groups would demonstrate within-subject decrease in SMA activation post-treatment during the fMRI finger tapping task (main effect of group: *F*(2, 42) = 0.11, *p* = .89, partial η² = .005; main effect of time: *F*(1, 42) = 0.28, *p* = .59, partial η² = .006; group x time interaction: *F*(2, 42) = 1.59, *p* = .21, partial η² = .07).

### SMA functional connectivity

Analyses confirmed the hypothesis that active TMS would alter within-subject RSFC connectivity of at least one ROI pair. Table 2 summarizes within-group RSFC changes (Cohen’s *d*) and associated effect sizes (η²) (spaghetti plots in Supplement). The largest effects were observed in the cTBS group for reduced connectivity between SMA and M1 Mouth 3 (cTBS η²=0.249, 1Hz η²=0.021, sham η²=0.018) and between putamen and M1 regions, including mouth (Mouth 1: cTBS η²=0.261, 1Hz η²=0.028, sham η²=0.158; Mouth 3: cTBS η²=0.298, 1Hz η²=0.003, sham η²=0.183) and hand areas (cTBS η²=0.265, 1Hz η²=0.031, sham η²=0.232). The reduction in SMA–M1 hand connectivity was more pronounced in the 1Hz group (η²=0.273) compared to the sham (η²<0.001) or cTBS (η²=0.046) groups.

**Table 2.**
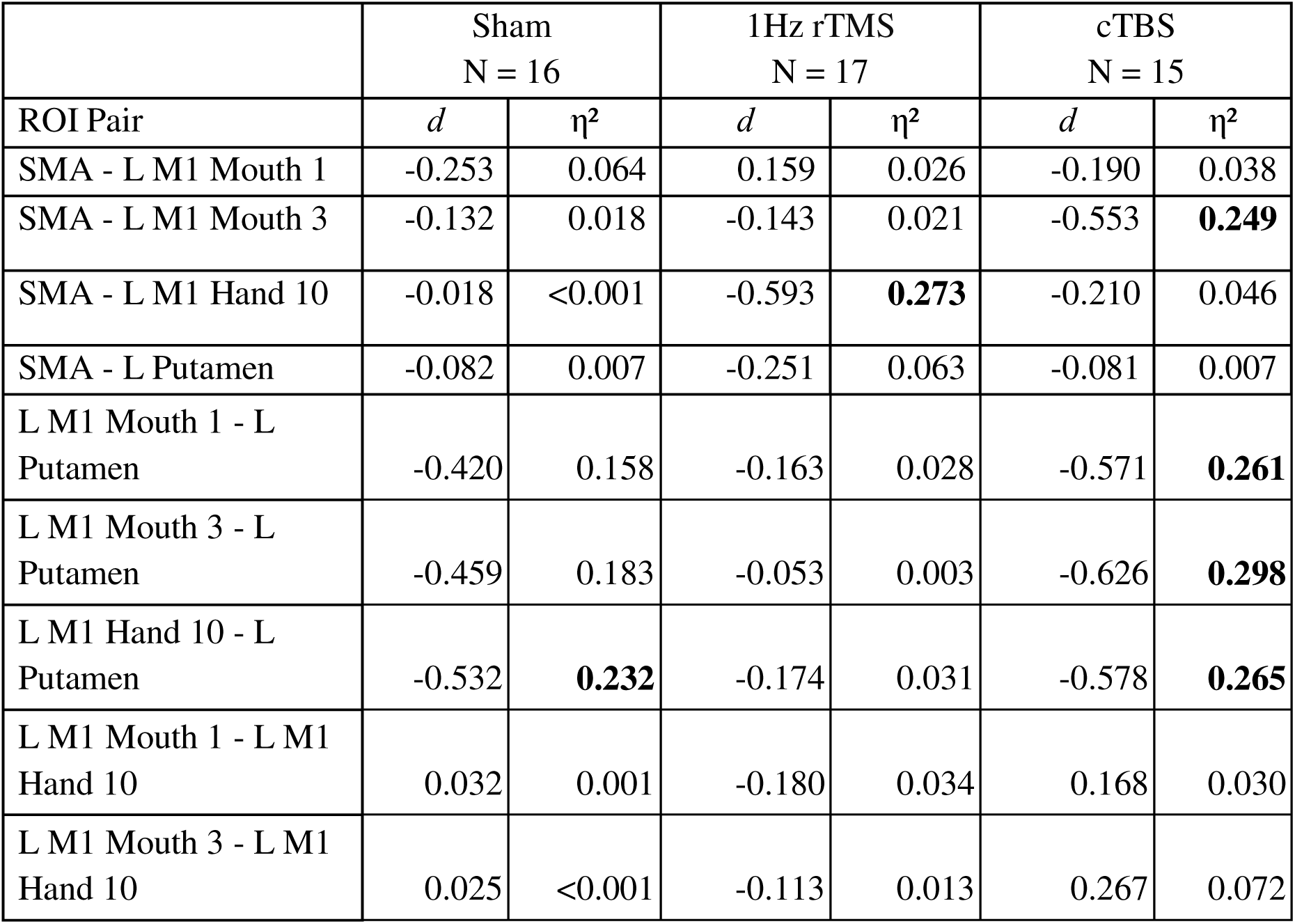
Mean RSFC differences (Cohen’s *d*) and effect sizes (η²) by group for ROI pairs.

The pattern of results suggests that CBIT alone (sham) had some effect on reducing cortical-subcortical connectivity (i.e., Putamen - M1 Hand, η²=0.232), but cTBS amplified this effect (i.e., greater Putamen - M1 reduction than sham). 1Hz appeared to blunt this effect (i.e., lower Putamen - M1 reduction than sham) and reduced cortical-cortical connectivity (SMA - M1).

### Behavioral target engagement

#### Tic frequency. Acute phase

Tic frequency during the TST Free-to-Tic condition was significantly reduced (β = -0.46, 95% CI: (-0.76, -0.15), p = .004) by a factor of 0.63 (37%, following exponentiation of the coefficient) following treatment. For the TST Suppression condition, tic frequency also significantly reduced (*β* = -0.49, 95% CI: (-0.87, -0.12), p = .012) by a factor of 0.61 (39%). For both conditions, there were no significant group differences across treatment or group-by-time interaction effects. Thus, all groups showed tic reduction and improved tic controllability.

#### Follow-up phase

Across all groups, tic frequency during the TST Free-to-Tic condition remained stable from post- to 1-month follow up (β = -0.20, 95% CI: (-0.60, -0.21), p = .333) but further reduced by 3-months follow-up (37% reduction vs. post-treatment, β = -0.46, 95% CI: (-0.86, -0.06), p = .025), suggesting continued improvement. For TST Suppression, we observed no significant changes in mean tic frequency across follow-ups, and neither group nor group by time effects were significant.

#### Premonitory urges. Acute phase

Mean urges decreased significantly in both TST Free-to-Tic (*β*= -1.16, 95% CI: (-1.93, -0.39), p = .004) and Suppression conditions (*β*= -1.71, 95% CI: (-2.50, -0.92), p < .001). We observed no significant group effects in either TST condition. However, we did observe a group-by-time interaction effect for the 1 Hz group in Free-to-Tic (*β* = 1.21, 95% CI: (1.13,2.29), p = .029) driven by a slight increase in urges in 1Hz rTMS and decreases in sham and cTBS groups (Supplement).

#### Follow-up phase

There were no significant differences in mean urges across the three-follow up time points in either TST condition. No overall group differences were observed to be significantly different among the three TMS groups, and there were no group-by-time interactions in either condition.

## DISCUSSION

This first test of TMS augmentation of behavior therapy for tics focused on establishing safety and feasibility, comparing 1Hz vs. cTBS protocols, and examining target engagement of SMA-mediated neurocircuitry and tic behaviors. Overall, results suggest cTBS is the preferred TMS regimen to pair with CBIT given its more favorable side effect profile and more robust engagement of tic-related circuitry vs. 1Hz rTMS.

Results support intervention feasibility, with a high completion rate, no TMS-related withdrawals, and high treatment satisfaction. TMS-related adverse events were mild and transient, consistent with prior literature in youth (47). A similar number of participants experienced a TMS-related AE in both active conditions, at higher rates than sham (∼70% vs. 50%). However, the number of sessions in which a treatment-related AE was reported was higher in 1Hz (27%) vs. cTBS and sham, which were comparable (sham = 11.3%, cTBS = 14.7%), suggesting that cTBS may be more tolerable for youth with tics than 1Hz. cTBS’s much briefer duration may contribute to tolerability.

In terms of neural target engagement, no group or time effects were seen for SMA task activation. It is possible that the bilateral finger-tapping task was not optimal for assessing TMS effects. Though the task robustly engages the motor system, it is externally cued, whereas SMA is preferentially recruited for internally-initiated movements (48). Therefore, TMS may have had limited effects on finger-tapping task activation, and a more complex, internally paced sequential tapping task may have been more likely to reveal changes. Such a task was used in Wu et al. (27), who observed decreased SMA activation in youth with tics following 8 cTBS sessions.

All participants in our study received active CBIT. RSFC data from the sham condition support the theory that CBIT promotes decreased hyperconnectivity of sensorimotor CSTC pathways (49). CBIT-related fMRI change has only been examined in one prior study of eight adults, which found a significant decrease in putamen activation during a visuospatial priming task post-CBIT (50). Presuming that our sham condition approximated the effect of CBIT monotherapy, our results converge with those of Deckersbach et al. (50) and suggest that CBIT may reduce functional connectivity between M1 and putamen, with small to medium effects.

In comparison, the cTBS group exhibited a potentiation and broadening of these cortical-striatal connectivity reductions, as evidenced by large effect sizes for all putamen to M1 subregion connections and a large effect for cortical-cortical decoupling between SMA and the M1 mouth area. The 1Hz group demonstrated essentially no change in cortico-striatal connectivity, instead showing only a localized cortical-cortical decoupling between SMA and the M1 hand area. Collectively, the RSFC results suggest that cTBS augmentation may amplify CBIT’s neural effects, whereas 1Hz may blunt them.

The finding that 1Hz potentially interfered with a neural mechanism of CBIT, while cTBS enhanced it, carries important implications for efforts to combine TMS with cognitive-behavioral therapies. Our results converge with prior research suggesting that 1Hz and cTBS are not functionally equivalent despite the heuristic that both are “inhibitory” (51). Modulatory effects may occur via distinct mechanisms (52) and depend on the functional connectivity profile of the targeted network (53). In M1, cTBS has been found to have more prominent and longer aftereffects vs. 1 Hz (54,55). Elsewhere in the cortex, these frequencies can induce “opposite” RSFC aftereffects. A direct comparison in SMA found that cTBS acutely reduced RSFC and 1Hz increased it (56), such that our results may reflect an excitatory effect of 1Hz that CBIT pairing effectively “neutralized.” Results also converge with research suggesting that some TMS protocols may interfere with psychotherapy mechanisms (57). Future studies should continue to use mechanistic, parametric tests to understand how different TMS protocols impact both disease-relevant and therapy-reliant circuitry. Moreover, as the SMA is one node within the larger action-mode network (58), it is important to consider that neuromodulation of this region is likely to affect function across the network.

Behavioral targets were measured using a direct observation paradigm, representing the first time the TST has been used in a clinical trial to measure change. All groups showed decreased tic frequency and controllability that further improved across the follow-up period, suggesting that all treatment arms had positive and lasting clinical effects. The only between-groups behavioral difference was decreased urges in sham and cTBS groups alongside increased urges in the 1Hz group across the acute phase and 3-month follow up. Results parallel the RSFC data pattern and converge with research implicating the SMA, and possibly the entire action-mode network, in the premonitory urge (59–62). Future studies should explore how TST-measured metrics of tic symptomology relate to neural measures and change with time and treatment, including whether treatments impact aspects of tics beyond frequency (e.g., intensity, temporal distribution).

This target engagement study was not designed to compare groups on clinical questionnaire measures, such that these were examined only descriptively. Importantly, we did not observe TMS-related iatrogenic clinical effects. Visual inspection of YGTSS scores suggest the possibility of more improvement in the cTBS group at 1-month follow-up. Phase 2 of this study is currently ongoing and powered to test whether changes in SMA RSFC mediate improvements on the YGTSS and TST. Given that enhanced outcomes in active TMS groups were not generally observed on behavioral and clinical measures, it will be important to establish whether TMS adds clinical benefits to CBIT in Phase 2.

Several limitations should be noted. The wide participant age range represented the typical window of treatment seeking, but this introduced variability in neural development that may have affected individual responses to TMS and CBIT (although average age was similar across treatment arms). A TMS monotherapy condition was not included, which limits our ability to compare results to prior TMS trials in tic disorders and to understand the neural effects of TMS alone. Finally, the treatment protocol duration and intensity was substantial for CBIT but brief for TMS. Prior tic research shows clinical benefit with longer stimulation regimens (29), such that results may differ with a greater “dose” of TMS. While this study shows TMS can engage SMA-related circuitry, more work is needed to confirm the clinical relevance of this circuitry and the optimal TMS dosing strategy for clinical improvement.

In conclusion, this early-stage trial demonstrates that cTBS to the SMA is a safe, feasible, and well-tolerated augmentation strategy for youth with tics undergoing CBIT. While 1Hz appeared to blunt the neural effects of behavior therapy, cTBS robustly engaged targeted sensorimotor circuitry, amplifying the reductions in cortico-striatal connectivity observed with CBIT alone. These findings support cTBS as the preferred TMS regimen for future trials powered to test clinical efficacy.

## Supporting information

Consort diagram

## Prior publications

Some data were reported previously in a published abstract (63).

## Author Contributions

CC: Conceptualization, Methodology, Writing-Original Draft, Writing - Review & Editing, Supervision, Project Administration, Funding acquisition. SK: Investigation, Project Administration. DG: Conceptualization, Methodology, Writing-Review & Editing. BM: Methodology, Software, Supervision, Writing-Review & Editing. CB: Investigation, Data Curation. KH: Investigation, Data Curation. BW: Investigation, Formal Analysis, Writing-Original Draft, Writing-Review & Editing. TH: Software, Visualization, Methodology, Formal Analysis. JA: Methodology, Investigation. MF: Formal Analysis. EC: Formal Analysis. KD: Data Curation, Formal Analysis, Writing-Original Draft. DS: Data Curation, Formal Analysis. MC: Methodology, Software, Supervision. CL: Writing-Reviewing & Editing. DR: Data Curation, Software, Validation. AO: Conceptualization, Methodology. SJ: Conceptualization, Investigation, Supervision. KL: Conceptualization, Methodology, Software, Supervision.

## Data availability

NIMH National Data Archive, Collection #3650, DOI: 10.15154/sc94-j082

## References

1. Centers for Disease Control and Prevention (CDC) (2009): Prevalence of diagnosed Tourette syndrome in persons aged 6-17 years - United States, 2007. MMWR Morb Mortal Wkly Rep 58: 581–585.

2. Conelea CA, Woods DW, Zinner SH, Budman C, Murphy T, Scahill LD, et al. (2011): Exploring the impact of chronic tic disorders on youth: results from the Tourette Syndrome Impact Survey. Child Psychiatry Hum Dev 42: 219–242.

3. Zinner SH, Conelea CA, Glew GM, Woods DW, Budman CL (2012): Peer victimization in youth with Tourette syndrome and other chronic tic disorders. Child Psychiatry Hum Dev 43: 124–136.

4. Piacentini J, Woods DW, Scahill L, Wilhelm S, Peterson AL, Chang S, et al. (2010): Behavior therapy for children with Tourette disorder: a randomized controlled trial. JAMA 303: 1929– 1937.

5. Wilhelm S, Peterson AL, Piacentini J, Woods DW, Deckersbach T, Sukhodolsky DG, et al. (2012): Randomized trial of behavior therapy for adults with Tourette syndrome. Arch Gen Psychiatry 69: 795–803.

6. McGuire JF, Piacentini J, Brennan EA, Lewin AB, Murphy TK, Small BJ, Storch EA (2014): A meta-analysis of behavior therapy for Tourette Syndrome. J Psychiatr Res 50: 106–112.

7. Woods DW, Piacentini J, Chang S, Deckersbach T, Ginsburg G, Peterson A, et al. (2008): Managing Tourette Syndrome: A Behavioral Intervention for Children and Adults Therapist Guide. Oxford University Press.

8. Pringsheim T, Okun MS, Müller-Vahl K, Martino D, Jankovic J, Cavanna AE, et al. (2019): Practice guideline recommendations summary: Treatment of tics in people with Tourette syndrome and chronic tic disorders. Neurology 92: 896–906.

9. Conelea CA, Wellen B, Woods DW, Greene DJ, Black KJ, Specht M, et al. (2018): Patterns and Predictors of Tic Suppressibility in Youth With Tic Disorders. Front Psychiatry 9: 188.

10. Wang Z, Maia TV, Marsh R, Colibazzi T, Gerber A, Peterson BS (2011): The neural circuits that generate tics in Tourette’s syndrome. Am J Psychiatry 168: 1326–1337.

11. Mink JW (2006): Neurobiology of basal ganglia and Tourette syndrome: basal ganglia circuits and thalamocortical outputs. Adv Neurol 99: 89–98.

12. Wardak C (2011): The Role of the Supplementary Motor Area in Inhibitory Control in Monkeys and Humans. J Neurosci 31: 5181–5183.

13. Bohlhalter S, Goldfine A, Matteson S, Garraux G, Hanakawa T, Kansaku K, et al. (2006): Neural correlates of tic generation in Tourette syndrome: an event-related functional MRI study. Brain 129: 2029–2037.

14. Hampson M, Tokoglu F, King RA, Constable RT, Leckman JF (2009): Brain areas coactivating with motor cortex during chronic motor tics and intentional movements. Biol Psychiatry 65: 594–599.

15. Stern E, Silbersweig DA, Chee KY, Holmes A, Robertson MM, Trimble M, et al. (2000): A functional neuroanatomy of tics in Tourette syndrome. Arch Gen Psychiatry 57: 741–748.

16. Franzkowiak S, Pollok B, Biermann-Ruben K, Südmeyer M, Paszek J, Thomalla G, et al. (2012): Motor-cortical interaction in Gilles de la Tourette syndrome. PLoS One 7: e27850.

17. Fox MD, Buckner RL, Liu H, Chakravarty MM, Lozano AM, Pascual-Leone A (2014): Resting-state networks link invasive and noninvasive brain stimulation across diverse psychiatric and neurological diseases. Proc Natl Acad Sci U S A 111: E4367–75.

18. Li Y, Yu L, Zhang H, Wang X, Cui Y, Li Y (2023): The severity and neural correlates of Premonitory Urge in Tourette syndrome: A systematic review and meta-analysis. J Integr Neurosci 22: 159.

19. Worbe Y, Malherbe C, Hartmann A, Pélégrini-Issac M, Messé A, Vidailhet M, et al. (2012): Functional immaturity of cortico-basal ganglia networks in Gilles de la Tourette syndrome. Brain 135: 1937–1946.

20. Fried I, Katz A, McCarthy G, Sass KJ, Williamson P, Spencer SS, Spencer DD (1991): Functional organization of human supplementary motor cortex studied by electrical stimulation. J Neurosci 11: 3656–3666.

21. Finis J, Enticott PG, Pollok B, Münchau A, Schnitzler A, Fitzgerald PB (2013): Repetitive transcranial magnetic stimulation of the supplementary motor area induces echophenomena. Cortex 49: 1978–1982.

22. Conelea CA, McLaughlin NCR (2019): Transcranial Magnetic Stimulation in Tourette Syndrome and Obsessive–Compulsive Disorder. Neurotechnology and Brain Stimulation in Pediatric Psychiatric and Neurodevelopmental Disorders. pp 189–215.

23. Le K, Liu L, Sun M, Hu L, Xiao N (2013): Transcranial magnetic stimulation at 1 Hertz improves clinical symptoms in children with Tourette syndrome for at least 6 months. J Clin Neurosci 20: 257–262.

24. Kwon HJ, Lim WS, Lim MH, Lee SJ, Hyun JK, Chae J-H, Paik KC (2011): 1-Hz low frequency repetitive transcranial magnetic stimulation in children with Tourette’s syndrome. Neurosci Lett 492: 1–4.

25. Kahl CK, Kirton A, Pringsheim T, Croarkin PE, Zewdie E, Swansburg R, et al. (2021): Bilateral transcranial magnetic stimulation of the supplementary motor area in children with Tourette syndrome. Dev Med Child Neurol 63: 808–815.

26. Leckman JF, Riddle MA, Hardin MT, Ort SI, Swartz KL, Stevenson J, Cohen DJ (1989): The Yale Global Tic Severity Scale: initial testing of a clinician-rated scale of tic severity. J Am Acad Child Adolesc Psychiatry 28: 566–573.

27. Wu SW, Maloney T, Gilbert DL, Dixon SG, Horn PS, Huddleston DA, et al. (2014): Functional MRI-navigated repetitive transcranial magnetic stimulation over supplementary motor area in chronic tic disorders. Brain Stimul 7: 212–218.

28. Bloch Y, Arad S, Levkovitz Y (2016): Deep TMS add-on treatment for intractable Tourette syndrome: A feasibility study. World J Biol Psychiatry 17: 557–561.

29. Landeros-Weisenberger A, Mantovani A, Motlagh MG, de Alvarenga PG, Katsovich L, Leckman JF, Lisanby SH (2015): Randomized Sham Controlled Double-blind Trial of Repetitive Transcranial Magnetic Stimulation for Adults With Severe Tourette Syndrome. Brain Stimul 8: 574–581.

30. Sack AT, Paneva J, Küthe T, Dijkstra E, Zwienenberg L, Arns M, Schuhmann T (2024): Target engagement and brain state dependence of transcranial magnetic stimulation: Implications for clinical practice. Biol Psychiatry 95: 536–544.

31. Conelea C, Greene D, Alexander J, Houlihan K, Hodapp S, Wellen B, et al. (2023): The CBIT+TMS Trial: study protocol for a two-phase randomized controlled trial testing neuromodulation to augment behavior therapy for youth with chronic tics. Res Sq. 10.21203/rs.3.rs-2949388/v1

32. American Psychiatric Association (2013): Diagnostic and Statistical Manual of Mental Disorders (DSM-5®). American Psychiatric Pub.

33. Wechsler D (n.d.): Wechsler Abbreviated Scale of intelligence. Retrieved July 20, 2023, from https://www.pearsonassessments.com/store/usassessments/en/Store/Professional-Assessments/Cognition-%26-Neuro/Wechsler-Abbreviated-Scale-of-Intelligence-%7C-Second-Edition/p/100000593.html

34. Woods DW, Piacentini J, Himle MB, Chang S (2005): Premonitory Urge for Tics Scale (PUTS): initial psychometric results and examination of the premonitory urge phenomenon in youths with Tic disorders. J Dev Behav Pediatr 26: 397–403.

35. Sheehan DV, Sheehan KH, Shytle RD, Janavs J, Bannon Y, Rogers JE, et al. (2010): Reliability and validity of the Mini International Neuropsychiatric Interview for Children and Adolescents (MINI-KID). J Clin Psychiatry 71: 313–326.

36. Sheehan DV, Lecrubier Y, Sheehan KH, Amorim P, Janavs J, Weiller E, et al. (1998): The Mini-International Neuropsychiatric Interview (M.I.N.I.): the development and validation of a structured diagnostic psychiatric interview for DSM-IV and ICD-10. J Clin Psychiatry 59 Suppl 20: 22–33;quiz 34–57.

37. Busner J, Targum SD (2007): The Clinical Global Impressions Scale: Applying a Research Tool in Clinical Practice. Psychiatry (Edgmont) 4: 28.

38. Krishnan C, Santos L, Peterson MD, Ehinger M (2015): Safety of noninvasive brain stimulation in children and adolescents. Brain Stimul 8: 76–87.

39. Attkisson CC, Zwick R (1982): The client satisfaction questionnaire. Psychometric properties and correlations with service utilization and psychotherapy outcome. Eval Program Plann 5: 233–237.

40. Woods DW, Himle MB (2004): Creating tic suppression: comparing the effects of verbal instruction to differential reinforcement. J Appl Behav Anal 37: 417–420.

41. DuBois M, Houlihan K, Raab B, Pryor A, Kellman M, Brinker M, et al. (2024): Quantifying tics: Best practices and design considerations for video-based tic coding in research. Behav Res Methods 56: 4073–4084.

42. Lab M (2020): TST Task Video. Retrieved July 29, 2026, from https://youtu.be/7-d_62nqKrk

43. The Adolescent Brain Cognitive Development (ABCD) study: Imaging acquisition across 21 sites (2018): Developmental Cognitive Neuroscience 32: 43–54.

44. Beynel L, Davis SW, Crowell CA, Hilbig SA, Lim W, Nguyen D, et al. (2019): Online repetitive transcranial magnetic stimulation during working memory in younger and older adults: A randomized within-subject comparison. PLoS One 14: e0213707.

45. blockrand: Randomization for Block Random Clinical Trials (n.d.): Comprehensive R Archive Network (CRAN). Retrieved February 18, 2025, from https://CRAN.R-project.org/package=blockrand

46. Seitzman BA, Gratton C, Marek S, Raut RV, Dosenbach NUF, Schlaggar BL, et al. (2020): A set of functionally-defined brain regions with improved representation of the subcortex and cerebellum. Neuroimage 206: 116290.

47. Allen CH, Kluger BM, Buard I (2017): Safety of Transcranial Magnetic Stimulation in Children: A Systematic Review of the Literature. Pediatr Neurol 68: 3–17.

48. Gowen E, Miall RC (2007): Differentiation between external and internal cuing: an fMRI study comparing tracing with drawing. Neuroimage 36: 396–410.

49. Eapen V, Črnčec R, Pick AX, Tsao O, Lai R, Lee J, Sowman PF (2022): Comprehensive behavioural intervention for tics: a neurophysiological intervention. J Integr Neurosci 21: 89.

50. Neural correlates of behavior therapy for Tourette s disorder (2014): Psychiatry Research: Neuroimaging 224: 269–274.

51. Beynel L, Powers JP, Appelbaum LG (2020): Effects of repetitive transcranial magnetic stimulation on resting-state connectivity: A systematic review. Neuroimage 211: 116596.

52. Di Lazzaro V, Dileone M, Pilato F, Capone F, Musumeci G, Ranieri F, et al. (2011): Modulation of motor cortex neuronal networks by rTMS: comparison of local and remote effects of six different protocols of stimulation. J Neurophysiol 105: 2150–2156.

53. Castrillon G, Sollmann N, Kurcyus K, Razi A, Krieg SM, Riedl V (2020): The physiological effects of noninvasive brain stimulation fundamentally differ across the human cortex. Sci Adv 6: eaay2739.

54. Huang Y-Z, Edwards MJ, Rounis E, Bhatia KP, Rothwell JC (2005): Theta burst stimulation of the human motor cortex. Neuron 45: 201–206.

55. Nyffeler T, Wurtz P, Lüscher H-R, Hess CW, Senn W, Pflugshaupt T, et al. (2006): Repetitive TMS over the human oculomotor cortex: comparison of 1-Hz and theta burst stimulation. Neurosci Lett 409: 57–60.

56. Dynamic aftereffects in supplementary motor network following inhibitory transcranial magnetic stimulation protocols (2017): NeuroImage 149: 285–294.

57. Fitzgerald PB, Jeff Daskalakis Z (2013): Repetitive Transcranial Magnetic Stimulation Treatment for Depressive Disorders: A Practical Guide. Springer Science & Business Media.

58. Dosenbach NUF, Raichle ME, Gordon EM (2025): The brain’s action-mode network. Nature Reviews Neuroscience 26: 158–168.

59. Greene DJ, Demeter DV, Nielsen AN (2025): Functional neuroanatomy of tics: A brain network perspective. Psychiatr Clin North Am 48: 31–44.

60. Worbe Y, Marrakchi-Kacem L, Lecomte S, Valabregue R, Poupon F, Guevara P, et al. (2015): Altered structural connectivity of cortico-striato-pallido-thalamic networks in Gilles de la Tourette syndrome. Brain 138: 472–482.

61. Wang Z, Maia TV, Marsh R, Colibazzi T, Gerber A, Peterson BS (2011): The neural circuits that generate tics in Tourette’s syndrome. Am J Psychiatry 168: 1326–1337.

62. Li Y, Yu L, Zhang H, Wang X, Cui Y, Li Y (2023): The severity and neural correlates of Premonitory Urge in Tourette syndrome: A systematic review and meta-analysis. J Integr Neurosci 22: 159.

63. ACNP 63rd Annual Meeting: Poster Abstracts P305-P608 (2024): Neuropsychopharmacology 49: 236–417.

