## Supplementary material for "1Hz vs. cTBS Transcranial Magnetic Stimulation to Augment Behavior Therapy for Tics: A Randomized Clinical Trial": Consort diagram

**Consort diagram: Phase 1 of the CBIT+TMS trial**

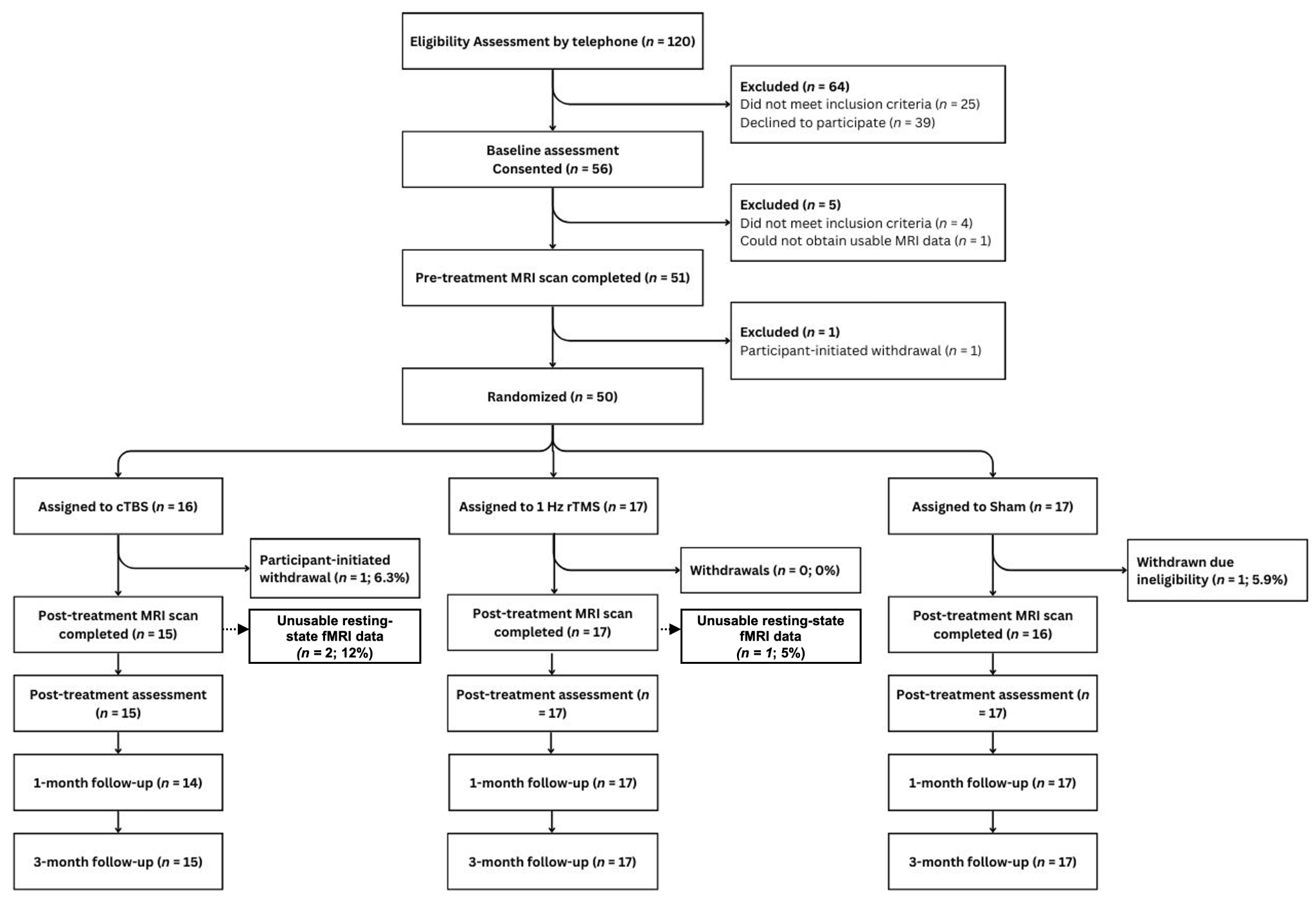

**Additional information about non-completers:** Non-completers (n = 2) included one cTBS participant who withdrew after session 1 due to disinterest in CBIT and another in sham who was withdrawn from TMS due to a status change that impacted their risk profile; this participant completed CBIT and all assessments except for the post-MRI.
