## Supplemental information for "1Hz vs. cTBS Transcranial Magnetic Stimulation to Augment Behavior Therapy for Tics: A Randomized Clinical Trial"

**Supplement**

1. **Power analysis**

Sample size was determined using *a priori* power analysis, which indicated that a sample of N = 48 has 80% power to detect 1) an effect size of at least η^2^=0.18 for within-subject change in SMA activation and at least η^2^=0.18 for within-subject change in RSFC and 2) an effect size of at least *d* = 0.90 for within-subject improvement in tic controllability.

2. **Patient and public involvement**

The Patient Advisory Board (PAB) of the Minnesota Tic and Compulsivity (MnTiC) Lab was involved in the design of the trial, including providing input in the questionnaire battery, recruitment and retention strategies, and interpretation of results. The PAB consists of 10 individuals with lived experience related to tic disorders and OCD. The PAB met monthly with lab staff and were compensated for their time and effort.

3. **Clinical assessment battery**

Tic severity was measured using the clinician-rated Yale Global Tic Severity Scale (YGTSS)(1). Ratings capture tic severity over the prior week and are summed to yield separate severity scores for Motor and Vocal Tics (range = 0-25) and a combined Total Tic score ranging from 0-50. Self-rated global premonitory urge intensity was measured with the Premonitory Urge for Tics Scale (PUTS; (2)), where scores range from 0-36, with higher scores reflecting greater urge intensity.

Psychiatric diagnoses were established with the MINI-7 KID (3) or Adult (4) and IQ with the Wechsler Abbreviated Scale of Intelligence (WASI; (5)). The Independent Evaluator’s (IE) impression of overall clinical improvement was captured with a Clinical-Global Impressions-Improvement (CGI-I (6)) scale adapted to guide ratings of tic-specific and global improvement. By convention, scores of 1 and 2 define a positive response. Safety/adverse events were assessed at every study visit using the TMS Adverse Effects Questionnaire (7) and the Ask Suicide-Screening Questions (8). Treatment satisfaction was measured at post-treatment with the Client Satisfaction Questionnaire-8 (9) by the participant and also a parent if a minor. Scores range from 4-32, with higher scores indicating greater satisfaction.

4. **Tic Suppression Task Reliability**

*Procedure:* Approximately 20% of videos in each context (47 videos total) were coded by two independent coders to confirm inter-rater reliability (IRR). Coder data were transformed into three-second bins. Bins containing one or more tics were labeled as “1” and bins with no tics present were labeled as “0”. Agreement was calculated as the percent of bins where both coders were aligned.

*Results:* Across all videos, the average agreement was 88% (range: 77% to 100%), consistent with established metrics in the field (10). Additionally, we calculated kappa agreement values. However, since kappa is greatly affected by variability, it tends to be low for videos where participants have few or no tics. Of the 47 double-coded videos, 39 (79%) had kappa values in the adequate range (kappa > .4). Of the eight videos where kappa was below .4, for 50%, at least one of the coders had coded two or fewer tics.

5. **MRI data: Additional information about acquisition and preprocessing.**

*Acquisition.* MR images were collected on a Siemens Prisma 3T scanner located at the Center for Magnetic Resonance Research (CMRR) at the University of Minnesota. All MRI data Structural, functional resting state and finger tapping task and EPI spin echo field map data) were acquired using the same pulse sequences used for the ABCD study (11).

1) Structural Scans: a) whole brain sagittal T1w MPRAGE using the motion-driven selective reacquisition of k-space to compensate for subject motion option (TR=2500ms, TE=2.9ms, TI=1070ms, 1.0mm isotropic voxel, flip angle = 8 degrees (7 min)) and b) whole brain sagittal T2 SPACE using the motion-driven selective reacquisition of k-space to compensate for subject motion option (TR=3200ms, TE=565ms, 1.0mm isotropic voxel, variable flip angle (6 min)).

2) Resting fMRI: Whole brain acquisition using the ABCD multi-band sequence (60 axial slices, 2.4mm isotropic voxel size, TR=800ms, TE=30ms, FOV=216mm, matrix=90x90, MB factor=6, 383 volumes (5 min)). During this scan participants were instructed to keep their eyes open and orient to a fixation cross. A total of 4 rest scans were typically acquired during each session.

3) Spin Echo EPI reverse phase encode scan pair: Whole brain acquisition using the ABCDspin echo sequence (60 axial slices, 2.4mm isotropic voxel size, TR=7030ms, TE=80ms, FOV=216mm, matrix=90x90, 1 volume pair (1 min))

4) Finger Tapping Task, for functional localization of SMA. Whole brain acquisition using HCP multi-band sequence (60 axial slices, 2.4mm isotropic voxel size, TR=800ms, TE=30ms, FOV=216mm, matrix=90x90, MB factor=6, 316 volume (4.25 min)). The task involved alternating 15sec blocks of simultaneous bilateral finger tapping and rest, cued by visual stimuli. This task isolates neural activity in motor planning and execution areas, including SMA, and is a highly reliable, well-established fMRI task used for motor mapping(12) and for TMS targeting of motor cortex and SMA.(13)

Motion considerations: To minimize motion artifacts, the participant’s head was stabilized using inflatable cushions, positioned to not contact the ears and inflated so the head was held snugly in the 32 channel head coil. This stabilization procedure minimized displacement of ear landmarks used to register participants to the TMS neuronavigation software.

Motion was monitored in real-time during all fMRI scans using Framewise Integrated Real-Time MRI Monitoring (FIRMM).(14,15) Acceptable volumes were based on a FD threshold of 0.2mm with respiration filter turned on. Scanner technicians were instructed to try to acquire a minimum of 10 “good” minutes of rsfMRI across the 4 runs, if necessary by acquiring a 5th resting scan if time allowed.

The final number of “usable” minutes was defined as the total number of minutes surviving post-hoc data cleaning with ≥5 contiguous frames of <0.2 mm FD (16). For inclusion in our resting-state fMRI analyses, we set a criterion of ≥6 minutes of FD-post-hoc volumes at both pre- and post-treatment scans. Three participants did not meet this threshold and were therefore not included in resting-state analyses (n = 2 in cTBS group, n = 1 in 1Hz group).

*Data processing.* The ABCD-BIDS HCP pipeline was used for MRI data processing.(17,18) This pipeline is based on the Human Connectome Project minimal processing pipeline.(19)

6. **TMS Targeting**

TMS targets were individually defined computationally using structural MRI, task fMRI, and finite electric field modeling data, as in Baynel et al. (17,18). This approach accounts for individual differences in physiology, which are prominent in developmental samples. Each participant’s pre-treatment structural MRI images (T1, T2) were input into SimNIBS to create individual finite element method (FEM) models (20–22) across 20 candidate locations in the SMA (using a digital 4x5 grid of points equally spaced on the scalp with 5mm intervals placed over a canonical SMA region of interest defined from the Harvard-Oxford cortical atlas (23) within MNI standard space). Coil orientation was fixed to point directly anterior and perpendicular to the scalp, based on prior literature, in consideration of operator ease and accuracy, and to constrain computational time. In parallel, finger tapping task fMRI functional maps were generated. BOLD signal for blocks of rest vs. tap within the SMA were compared using a general linear model and statistical parameter maps (SPMs). In the final step, pairwise correlations were calculated for the generated SMA-constrained 20 electric field maps and task fMRI SPMs. The grid location and orientation (x, y, z coordinates) with the largest correlation was selected as the TMS target. This target was input into a stereotaxic neuronavigation system (BrainSight 2.3.5, Rogue Research, Montreal, Quebec, Canada) for TMS delivery. Targeting was adjusted slightly due to pipeline issues for five participants and due to tolerability for one participant (detailed in Supplement 6).

Figure S2. Schematic overview of the TMS stimulation targeting pipeline.

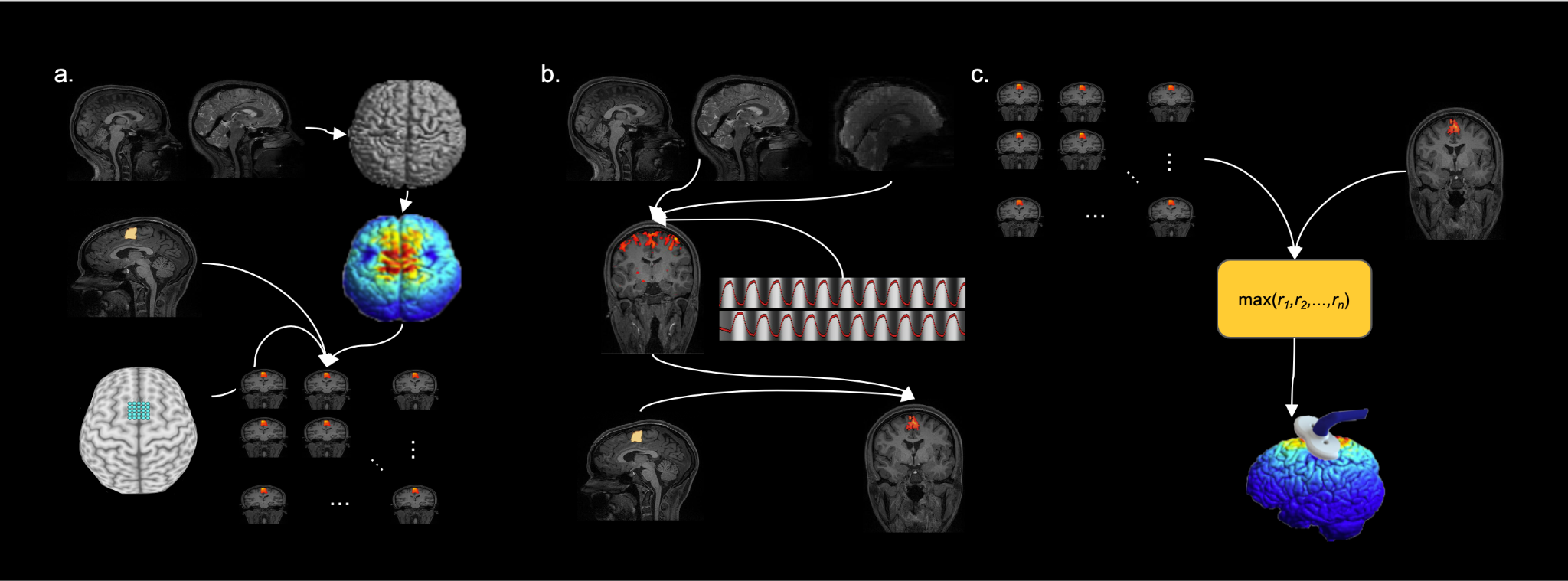

The pipeline is split into three separate stages: (**a)** simulated induced electric current map generation, (**b)** task fMRI functional map generation, and (**c)** selection of stimulation target. (**a)** Individual T1-weighted and T2-weighted anatomical image pairs (upper left) were used to produce a tetrahedral head mesh (upper right). Electric fields were simulated for each of the 20 native space coordinates and masked based on the SMA parcel (bottom right). Representative induced electric field simulation (middle left). (**b)** Second, anatomical and BOLD images (top) and finger tapping task timing (middle right) a statistical parametric map (SPM) was produced representing finger tapping functional activation. A mask of SMA (bottom left) was used to extract finger tapping SPM values only within SMA (bottom right). (**c)** Third, the finger tapping SPM (upper right) and simulated electric fields (upper left) for the 20 coordinates were correlated pairwise (middle). The coordinate corresponding to the maximum correlation was chosen as the stimulation target (middle, bottom).

7. **Adjustments to TMS targets.**

*Adjustments due to technical issues with the targeting pipeline* were made for five participants.

For 3 of these participants, activation differences between the tap and rest blocks of the motor fMRI task were weak. Based on staff observations during the scans, weak activations appeared to be related to factors such as tic-related motion or movement inhibition attempts during “rest” blocks. For these participants, TMS targets were selected based on the SimNIBS-generated highest e-field value in SMA. For 2 of these participants, we encountered errors within SimNIBS. In one instance, SimNIBS failed to extract the surface mesh into volume space. One was related to a file permissions issue that could not be resolved in time for the start of stimulation. In both of these cases, we calculated the center of gravity voxel within SMA based on the task fMRI activation. The coordinate on the 4x5 grid closest to this voxel was selected as the TMS target.

*Adjustments due to tolerability issues* were made for one participant in the 1Hz group, who reported localized pain during delivery of TMS pulses at session 2. Tolerability was improved with a minor adjustment in coil placement (1mm diagonal to the initial target in the anterior-lateral direction) and reduction in stimulation intensity to 80% of RMT)

8. **Target locations in SMA by parcel**

Multiple Steitzman et al. (24) atlas parcels fall within the SMA. As noted in the main manuscript, the parcel containing the individual’s TMS target was used for the SMA ROI in connectivity analyses (see Fig. 1). The table below details the parcels that were used for the SMA ROI for the n = 45 participants represented in the RSFC analyses:

L_CinguloOperc_5: 19

L_VentralAttn_1: 16

R_CinguloOperc_26: 5

R_CinguloOperc_27: 5

9. **CBIT Fidelity**

*Procedure*: 55 CBIT sessions were randomly selected for fidelity rating, representing 11% of the total number of delivered sessions. Fidelity raters were three doctoral-level clinical psychologists with expertise in CBIT; none acted as therapists in the study. Raters reviewed session video recordings and transcriptions and completed fidelity rating forms used in prior CBIT research (25). These forms focus on capturing the amount of the session spent on prescribed vs. proscribed elements of the session according to the study CBIT manual. Raters also judged the overall fidelity of the session on a likert-type scale ranging from 1 (Poor Fidelity) to 7 (Good Fidelity).

*Results*: Overall fidelity ratings had a mean of 6.88 (SD = 0.45, range = 3-7), representing high overall fidelity of sessions to the CBIT manual. Ratings of adherence are reported in the tables below.

Table S7.a. Overall Adherence to the Specific Session (N=55).

|  | % (N) |
| --- | --- |
| 0: <25% of the session was spent on elements scheduled in the manual for this session | 0% (0) |
| 1: 25% - 75% of the session was spent on elements scheduled in the manual for this session | 10.9% (6) |
| 2: 75% - 100% of the session was spent on elements scheduled in the manual for this session | 89.09% (49) |

Table S7.b. Overall Adherence to CBIT Components (N=55)

|  | % (N) |
| --- | --- |
| 0: <25% of the session was spent on elements contained in the CBIT protocol | 7.27% (4) |
| 1: 25% - 75% of the session was spent on elements contained in the CBIT protocol | 7.27% (4) |
| 2: 75% - 100% of the session was spent on elements contained in the CBIT protocol | 85.45% (47) |

Table S7.c. Use of non-CBIT CBT components (N=55)

|  | % (N) |
| --- | --- |
| 0: <25% of the session was spent on CBT elements NOT contained in the CBIT protocol | 98.18% (54) |
| 1: 25% - 75% of the session was spent on CBT elements NOT contained in the CBIT protocol | 1.82% (1) |
| 2: 75% - 100% of the session was spent on CBT elements NOT contained in the CBIT protocol | 0% (0) |

Table S7.d. Use of non-CBIT supportive/non-specific components (N=55)

|  | % (N) |
| --- | --- |
| 0: <25% of the session was spent on non-specific elements NOT contained in the CBIT protocol | 100% (55) |
| 1: 25% - 75% of the session was spent on non-specific elements NOT contained in the CBIT protocol | 0% (0) |
| 2: 75% - 100% of the session was spent on non-specific elements NOT contained in the CBIT protocol | 0% (0) |

10. **TMS Blinding**

*Procedure:* A questionnaire assessing beliefs about TMS treatment assignment was administered to participants and parents (for minors) at post-treatment, 1-month follow-up, and 3-month follow-up. The IE, CBIT therapist, and TMS operator completed the questionnaire at post-treatment only. The questionnaire consisted of two items: 1) “What type of stimulation do you believe you received?” and 2) “How certain are you of the type of stimulation you received?” Participants could optionally share the reason for their choice in an open-text box. Items 1 and 2 were analyzed using descriptive statistics.

*Results:* Descriptive data for each reporter group are presented in the tables below. Overall, results show that all reporter groups guessed TMS assignment at levels below chance. Notably, certainty ratings indicate that very few participants and parents believed they received sham stimulation, and the most commonly endorsed rating by all was “Don’t know.” Cumulatively, this pattern of results indicates that masking procedures were adequate.

Table S8.a: Participant Responses to TMS Masking Questionnaire

| **Characteristic** | **Post**, N = 48*^1^* | **1Month**, N = 48*^1^* | **3Month**, N = 48*^1^* |
| --- | --- | --- | --- |
| Correct_guess | 21 (44%) | 21 (46%) | 22 (46%) |
| Unknown | 0 | 2 | 0 |

| **Characteristic** | **Post**,  N = 48*^1^* | **1Month**, N = 48*^1^* | **3Month**, N = 48*^1^* |
| --- | --- | --- | --- |
| *Certainty Rating* |  |  |  |
| Strongly believe the stimulation was active | 5 (10%) | 7 (15%) | 8 (17%) |
| Somewhat believe the stimulation was active | 16 (33%) | 13 (28%) | 13 (27%) |
| Don’t know | 21 (44%) | 16 (35%) | 19 (40%) |
| Somewhat believe the stimulation was sham | 6 (13%) | 9 (20%) | 8 (17%) |
| Strongly believe the stimulation was sham | 0 (0%) | 1 (2.2%) | 0 (0%) |
| Unknown | 0 | 2 | 0 |
| *^1^* n (%) | | | |

Table S8.b: Parent Responses to TMS Masking Questionnaire

| **Characteristic** | **Post**, N = 48*^1^* | **1Month**, N = 48*^1^* | **3Month**, N = 48*^1^* |
| --- | --- | --- | --- |
| Correct_guess | 11 (31%) | 12 (34%) | 11 (34%) |
| Unknown* | 12 | 13 | 16 |

| **Characteristic** | **Post**, N = 48*^1^* | **1Month**, N = 48*^1^* | **3Month**, N = 48*^1^* |
| --- | --- | --- | --- |
| *Certainty Rating* |  |  |  |
| Strongly believe the stimulation was active | 8 (22%) | 6 (17%) | 7 (22%) |
| Somewhat believe the stimulation was active | 3 (8.3%) | 4 (11%) | 8 (25%) |
| Don’t know | 19 (53%) | 19 (54%) | 11 (34%) |
| Somewhat believe the stimulation was sham | 5 (14%) | 5 (14%) | 5 (16%) |
| Strongly believe the stimulation was sham | 1 (2.8%) | 1 (2.9%) | 1 (3.1%) |
| Unknown* | 12 | 13 | 16 |
| *^1^* n (%)  *This questionnaire was not collected for adult participants. | | | |

Table S8.c: Independent Evaluator (IE) Responses to TMS Masking Questionnaire

| **Characteristic** | **Post**, N = 48*^1^* |
| --- | --- |
| Correct_guess | 5 (10%) |
| *^1^* n (%) | |

| **Characteristic** | **Post**, N = 48*^1^* |
| --- | --- |
| *Certainty Rating* |  |
| Strongly believe the stimulation was active | 2 (4.2%) |
| Somewhat believe the stimulation was active | 2 (4.2%) |
| Don’t know | 42 (88%) |
| Somewhat believe the stimulation was sham | 2 (4.2%) |
| Strongly believe the stimulation was sham | 0 (0%) |
| *^1^* n (%) |  |

Table S8.c: CBIT Therapist Responses to TMS Masking Questionnaire

| **Characteristic** | **Post**, N = 48*^1^* |
| --- | --- |
| Correct_guess | 15 (31%) |
| *^1^* n (%) | |

| **Characteristic** | **Post**, N = 48*^1^* |
| --- | --- |
| *Certainty Rating* |  |
| Strongly believe the stimulation was active | 3 (6.3%) |
| Don’t know | 12 (25%) |
| Somewhat believe the stimulation was sham | 21 (44%) |
| Strongly believe the stimulation was sham | 12 (25%) |
| *^1^* n (%) | |

11. **Adverse Events**

Table S9.a: Acute Phase “Person-Level” AEs: Participants who Experienced One or More Adverse Events Judged to be Possibly or Likely Related to TMS Across Groups

| Adverse Event Category | cTBS (*n* = 15) | | 1 Hz rTMS (*n* = 17) | | Sham (*n* = 16) | |
| --- | --- | --- | --- | --- | --- | --- |
|  | Count | % (out of 15) | Count | % (out of 17) | Count | % (out of 16) |
| Headache | 6 | 40.0 | 7 | 41.2 | 7 | 43.8 |
| Muscle Twitch | 0 | 0.0 | 7 | 41.2 | 2 | 12.5 |
| Sleep Disturbance | 3 | 20.0 | 4 | 23.5 | 1 | 6.3 |
| Other | 1 | 6.7 | 2 | 11.8 | 2 | 12.5 |
| Mood Change | 1 | 6.7 | 2 | 11.8 | 0 | 0.0 |
| Pain | 1 | 6.7 | 1 | 5.9 | 2 | 12.5 |
| Nausea | 1 | 6.7 | 1 | 5.9 | 0 | 0.0 |
| Scalp Discomfort | 0 | 0.0 | 2 | 11.8 | 0 | 0.0 |
| Concentration Difficulties | 1 | 6.7 | 1 | 5.9 | 0 | 0.0 |
| Hearing Difficulties | 1 | 6.7 | 0 | 0.0 | 0 | 0.0 |
| Memory Difficulties | 0 | 0.0 | 0 | 0.0 | 0 | 0.0 |
| Syncope | 0 | 0.0 | 0 | 0.0 | 0 | 0.0 |
| Seizure | 0 | 0.0 | 0 | 0.0 | 0 | 0.0 |
| Any Adverse Event | 11 | 73.3 | 12 | 70.6 | 8 | 50 |

Table S9.b.: Follow-Up Phase “Person-Level” AEs: Participants who Experienced One or More Adverse Events Judged to be Possibly or Likely Related to TMS Across Groups in the Follow-Up Period

| Adverse Event Category | cTBS (*n* = 15) | | 1 Hz rTMS (*n* = 17) | | Sham (*n* = 16) | |
| --- | --- | --- | --- | --- | --- | --- |
|  | Count | % (out of 15) | Count | % (out of 17) | Count | % (out of 16) |
| Headache | 0 | 0 | 2 | 11.8 | 1 | 6.3 |
| Muscle Twitch | 1 | 6.7 | 1 | 5.9 | 1 | 6.3 |
| Sleep Disturbance | 0 | 0 | 0 | 0 | 0 | 0 |
| Other | 0 | 0 | 0 | 0 | 0 | 0 |
| Mood Change | 0 | 0 | 0 | 0 | 0 | 0 |
| Pain | 0 | 0 | 1 | 5.9 | 0 | 0 |
| Nausea | 0 | 0 | 0 | 0 | 0 | 0 |
| Scalp Discomfort | 0 | 0 | 0 | 0 | 0 | 0 |
| Concentration Difficulties | 0 | 0 | 0 | 0 | 1 | 6.3 |
| Hearing Difficulties | 0 | 0 | 1 | 5.9 | 0 | 0 |
| Memory Difficulties | 0 | 0 | 0 | 0 | 0 | 0 |
| Syncope | 0 | 0 | 0 | 0 | 0 | 0 |
| Seizure | 0 | 0 | 0 | 0 | 0 | 0 |
| Any Adverse Event | 1 | 6.7 | 5 | 23.5 | 3 | 18.8 |

Table S9.c: Acute Phase “Session-Level” AEs: Adverse Events Judged to be Possibly or Likely Related to TMS Across Groups

| Adverse Event Category | cTBS (*n* = 15) | | 1 Hz rTMS (*n* = 17) | | Sham (*n* = 16) | |
| --- | --- | --- | --- | --- | --- | --- |
|  | Count | %/Rate (out of 150 TMS sessions) | Count | Rate (out of 170 TMS sessions) | Count | Rate (out of 160 TMS sessions) |
| Headache | 10 | 6.7 | 21 | 12.4 | 9 | 5.6 |
| Muscle Twitch | 0 | 0.0 | 8 | 4.7 | 2 | 1.3 |
| Sleep Disturbance | 3 | 2.0 | 5 | 2.9 | 1 | 0.6 |
| Other | 1 | 0.7 | 2 | 1.2 | 4 | 2.5 |
| Mood Change | 2 | 1.3 | 4 | 2.4 | 0 | 0.0 |
| Pain | 2 | 1.3 | 2 | 1.2 | 2 | 1.3 |
| Nausea | 2 | 1.3 | 1 | 0.6 | 0 | 0.0 |
| Scalp Discomfort | 0 | 0.0 | 2 | 1.2 | 0 | 0.0 |
| Concentration Difficulties | 1 | 0.7 | 1 | 0.6 | 0 | 0.0 |
| Hearing Difficulties | 1 | 0.7 | 0 | 0.0 | 0 | 0.0 |
| Memory Difficulties | 0 | 0.0 | 0 | 0.0 | 0 | 0.0 |
| Syncope | 0 | 0.0 | 0 | 0.0 | 0 | 0.0 |
| Seizure | 0 | 0.0 | 0 | 0.0 | 0 | 0.0 |
| Total Adverse Events | 22 | 14.7 | 46 | 27.1 | 18 | 11.3 |

Table S9.d: Follow-Up Phase “Session-Level” AEs: Adverse Events Judged to be Possibly or Likely Related to TMS Across Groups During the Follow-Up Period

| Adverse Event Category | cTBS (*n* = 15) | | 1 Hz rTMS (*n* = 17) | | Sham (*n* = 16) | |
| --- | --- | --- | --- | --- | --- | --- |
|  | Count | %/Rate (out of 30 follow-up visits) | Count | Rate (out of 34 follow-up visits) | Count | Rate (out of 32 follow-up visits) |
| Headache | 0 | 0 | 2 | 5.9 | 1 | 3.1 |
| Muscle Twitch | 1 | 3.3 | 1 | 2.9 | 2 | 6.3 |
| Sleep Disturbance | 0 | 0 | 0 | 0 | 0 | 0 |
| Other | 0 | 0 | 0 | 0 | 0 | 0 |
| Mood Change | 0 | 0 | 0 | 0 | 0 | 0 |
| Pain | 0 | 0 | 1 | 2.9 | 0 | 0 |
| Nausea | 0 | 0 | 0 | 0 | 0 | 0 |
| Scalp Discomfort | 0 | 0 | 0 | 0 | 0 | 0 |
| Concentration Difficulties | 0 | 0 | 0 | 0 | 1 | 3.1 |
| Hearing Difficulties | 0 | 0 | 1 | 2.9 | 0 | 0 |
| Memory Difficulties | 0 | 0 | 0 | 0 | 0 | 0 |
| Syncope | 0 | 0 | 0 | 0 | 0 | 0 |
| Seizure | 0 | 0 | 0 | 0 | 0 | 0 |
| Total Adverse Events | 1 | 3.3 | 5 | 14.7 | 4 | 12.5 |

12. **RSFC connectivity spaghetti plots.**

**Figure S3.** Plots of within-subject RSFC connectivity change for each ROI pair, by TMS stimulation group. “Target” reflects the SMA parcel in which an individual’s TMS target fell.

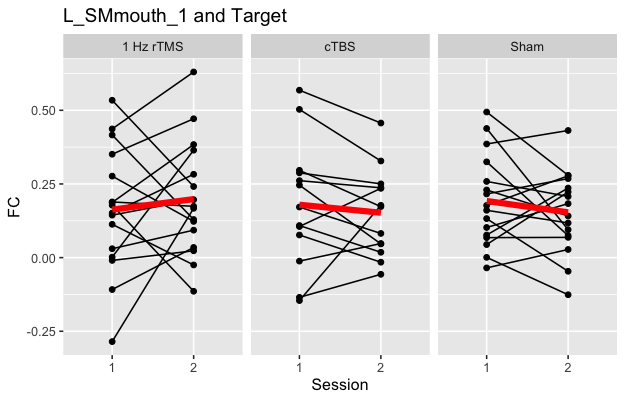

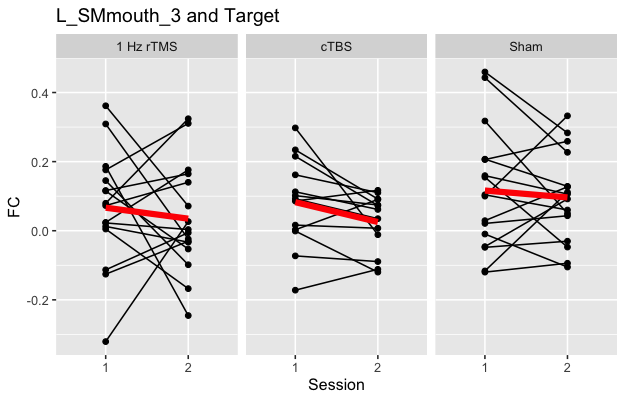

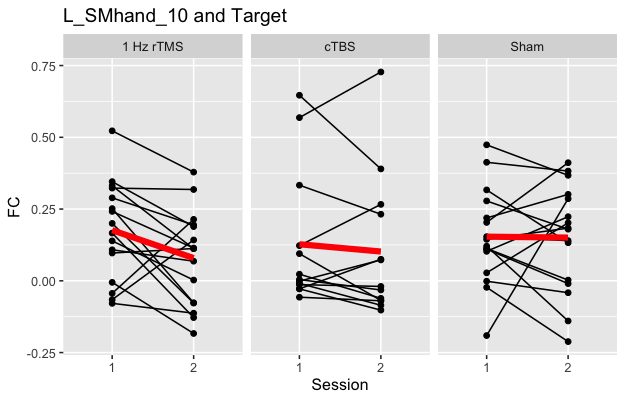

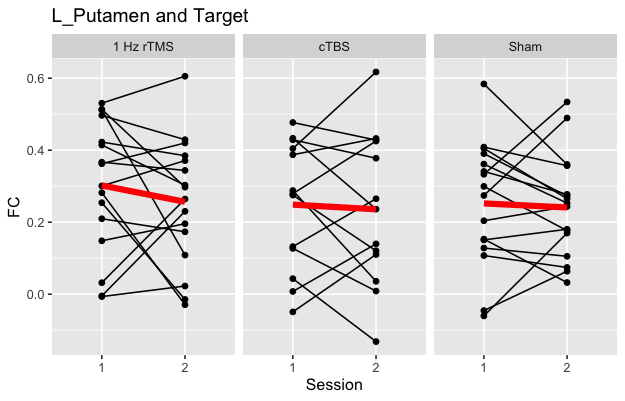

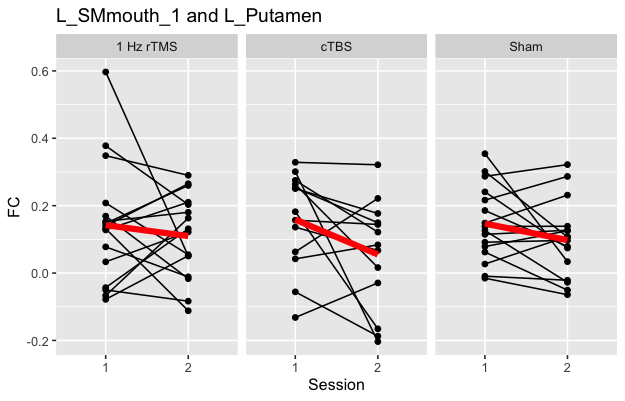

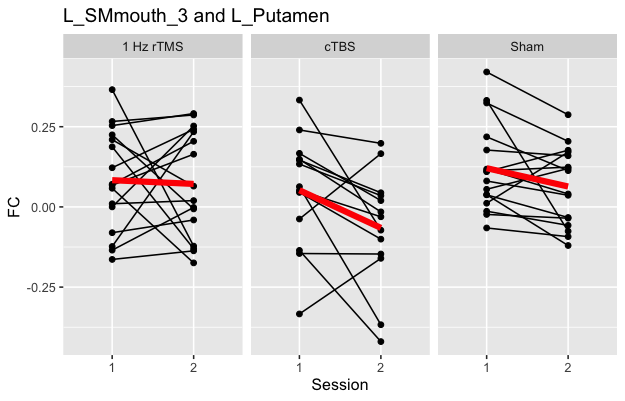

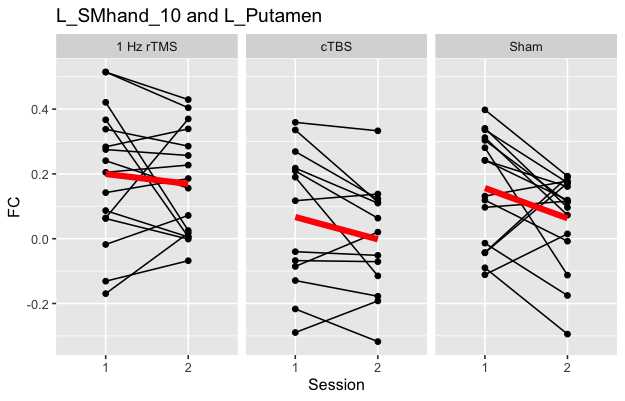

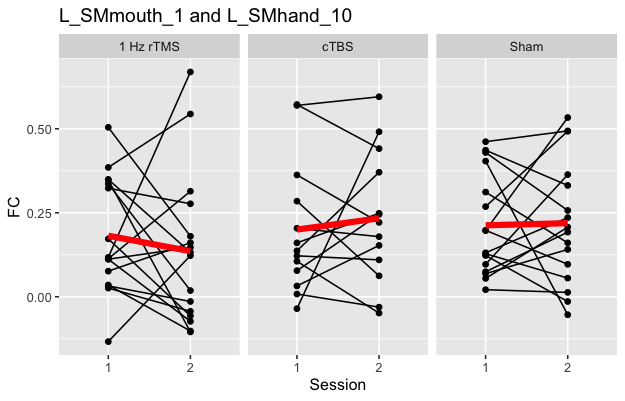

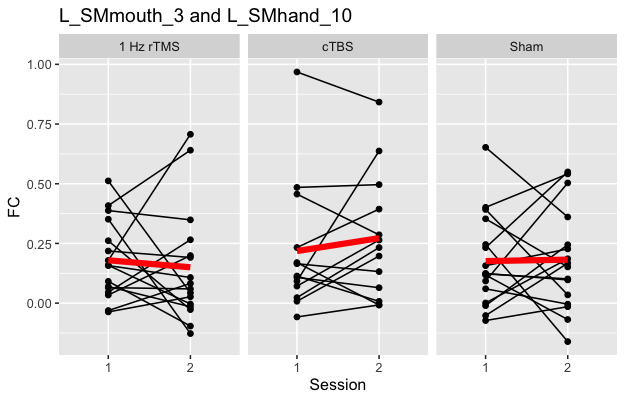

13. **Interaction plot for groupwise contrast of premonitory urge ratings during Tic Suppression Task from pre- to post-treatment.**

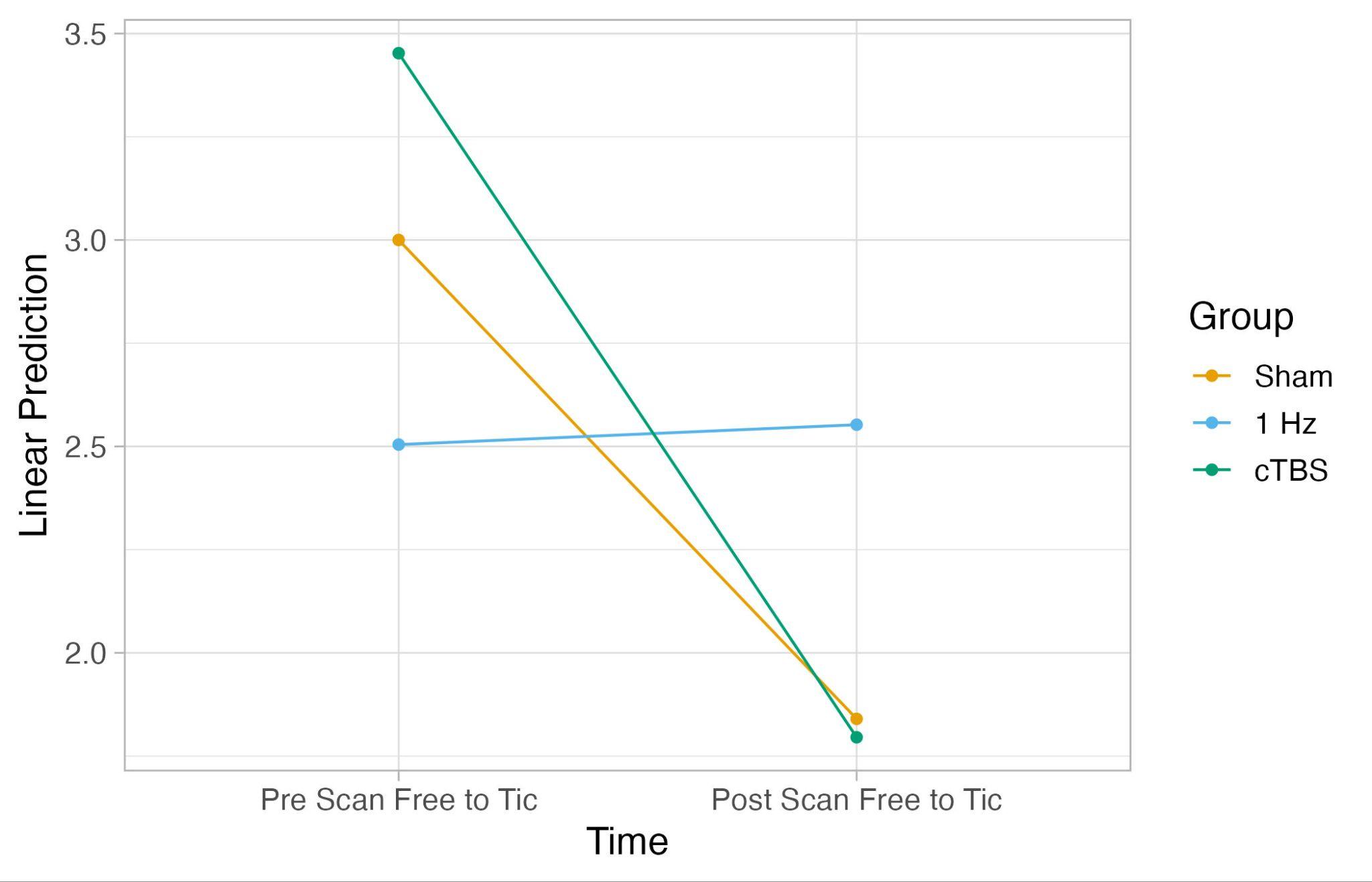

A follow-up comparison of the contrasts revealed that the two time points (pre- versus post-scan) were significantly different from each other in the Sham and cTBS groups, but not in the 1Hz group.

| **Group** | **Estimate (Pre-Scan - Post Scan Contrast)** | **SE** | **df** | **t** | ***p*-value** |
| --- | --- | --- | --- | --- | --- |
| Sham | 1.160 | 0.381 | 43.0 | 3.044 | 0.004 |
| 1 Hz | -0.048 | 0.379 | 43.9 | -0.127 | 0.900 |
| cTBS | 1.657 | 0.383 | 42.1 | 4.324 | <0.0001 |

14. **Exploratory examination of clinical assessment measures**

To identify potential negative effects of TMS on clinical functioning, we descriptively explored YGTSS and PUTS data patterns across timepoints. CGI-I scores were examined at post-treatment and 1- and 3-month follow-ups.

**Treatment of missing data.** For clinical/behavioral data, one randomized participant missed a full assessment (1-month follow-up). Minimal (<3%) missing item-level data in the PUTS and CSQ was imputed using the person-specific mean from the non-missing items at the according time point.

Group-level YGTSS scores were somewhat uneven at baseline (sham m = 26, 1Hz rTMS m = 24, cTBS m = 20, *p* = 0.08), making direct comparisons difficult. However, all groups had lower mean YGTSS scores at post- and follow-ups compared to baseline (*p* <.05). PUTS scores were relatively stable for all groups, although average scores showed an increasing pattern over time through 3-month follow-up in the 1Hz rTMS group. On the CGI-I, IEs rated sham participants “improved” somewhat more frequently than active TMS participants, except at the 1-month follow-up where more cTBS participants were rated as improved. Collectively, these results indicate that TMS augmentation did not seem to be associated with iatrogenic clinical effects.

**YGTSS**

| Characteristic | Pre | | | Post | | | 1Month | | | 3Month | | |
| --- | --- | --- | --- | --- | --- | --- | --- | --- | --- | --- | --- | --- |
|  | Sham*^1^* | 1 Hz*^1^* | cTBS*^1^* | Sham*^1^* | 1 Hz*^1^* | cTBS*^1^* | Sham*^1^* | 1 Hz*^1^* | cTBS*^1^* | Sham*^1^* | 1 Hz*^1^* | cTBS*^1^* |
| **Total Severity** | 26 (25, 31) | 24 (20, 29) | 20 (18, 24) | 21.0 (18.0, 24.5) | 22.0 (19.0, 25.0) | 19.0 (16.0, 21.0) | 21.5 (15.5, 23.5) | 21.0 (16.0, 24.0) | 12.5 (9.0, 19.0) | 20.0 (16.0, 21.5) | 19.0 (17.0, 27.0) | 18.0 (13.0, 20.0) |
| Unknown |  |  |  |  |  |  | 0 | 0 | 1 |  |  |  |
| *^1^* Median (IQR) | | | | | | | | | | | | |

**PUTS**

| **Characteristic** | **Pre** | | | **Post** | | | **1Month** | | | **3Month** | | |
| --- | --- | --- | --- | --- | --- | --- | --- | --- | --- | --- | --- | --- |
|  | **Sham***^1^* | **1 Hz***^1^* | **cTBS***^1^* | **Sham***^1^* | **1 Hz***^1^* | **cTBS***^1^* | **Sham***^1^* | **1 Hz***^1^* | **cTBS***^1^* | **Sham***^1^* | **1 Hz***^1^* | **cTBS***^1^* |
| PUTS Score  Pre-Tx | 14 (8, 19) | 11 (6,17) | 12 (7, 19) | 13 (6, 16) | 13 (6, 18) | 13 (6, 16) | 12 (6,17) | 14 (4, 17) | 13 (10, 15) | 14 (9, 18) | 17 (6, 18) | 11 (9, 15) |
| Unknown |  |  |  |  |  |  | 0 | 0 | 1 |  |  |  |
| *^1^* Median (Q1, Q3) | | | | | | | | | | | | |

**CGI: IE rating of tic-specific improvement.** Note that “Improvement” corresponds to responses of “Very much better” or “Much better”, whereas “Little to no Improvement” corresponds to any other response (“A little better”, “No change”, “A little worse”, “Much worse”, “Very much worse”):

|  | **Pre** | | | **Post** | | | **1Month** | | | **3Month** | | |
| --- | --- | --- | --- | --- | --- | --- | --- | --- | --- | --- | --- | --- |
| **Characteristic** | **Sham***^1^* | **1 Hz***^1^* | **cTBS***^1^* | **Sham***^1^* | **1 Hz***^1^* | **cTBS***^1^* | **Sham***^1^* | **1 Hz***^1^* | **cTBS***^1^* | **Sham***^1^* | **1 Hz***^1^* | **cTBS***^1^* |
| Improvement |  |  |  |  |  |  |  |  |  |  |  |  |
| Improvement | 0 (NA%) | 0 (NA%) | 0 (NA%) | 9 (56%) | 6 (35%) | 5 (33%) | 7 (44%) | 8 (47%) | 9 (64%) | 10 (63%) | 7 (41%) | 7 (47%) |
| Little to no Improvement | 0 (NA%) | 0 (NA%) | 0 (NA%) | 7 (44%) | 11 (65%) | 10 (67%) | 9 (56%) | 9 (53%) | 5 (36%) | 6 (38%) | 10 (59%) | 8 (53%) |
| Unknown | 16 | 17 | 15 |  |  |  | 0 | 0 | 1 |  |  |  |
| *^1^*n (%) | | | | | | | | | | | | |

**CGI: IE rating of** **global improvement**. Note that “Improvement” corresponds to responses of “Very much better” or “Much better”, whereas “Little to no Improvement” corresponds to any other response (“A little better”, “No change”, “A little worse”, “Much worse”, “Very much worse”):

|  | **Pre** | | | **Post** | | | **1Month** | | | **3Month** | | |
| --- | --- | --- | --- | --- | --- | --- | --- | --- | --- | --- | --- | --- |
| **Characteristic** | **Sham***^1^* | **1 Hz***^1^* | **cTBS***^1^* | **Sham***^1^* | **1 Hz***^1^* | **cTBS***^1^* | **Sham***^1^* | **1 Hz***^1^* | **cTBS***^1^* | **Sham***^1^* | **1 Hz***^1^* | **cTBS***^1^* |
| Improvement |  |  |  |  |  |  |  |  |  |  |  |  |
| Improvement | 0 (NA%) | 0 (NA%) | 0 (NA%) | 7 (44%) | 6 (35%) | 3 (20%) | 8 (50%) | 8 (47%) | 5 (36%) | 7 (44%) | 5 (29%) | 5 (33%) |
| Little to no Improvement | 0 (NA%) | 0 (NA%) | 0 (NA%) | 9 (56%) | 11 (65%) | 12 (80%) | 8 (50%) | 9 (53%) | 9 (64%) | 9 (56%) | 12 (71%) | 10 (67%) |
| Unknown | 16 | 17 | 15 |  |  |  | 0 | 0 | 1 |  |  |  |
| *^1^*n (%) | | | | | | | | | | | | |

**References**

1. Leckman JF, Riddle MA, Hardin MT, Ort SI, Swartz KL, Stevenson J, Cohen DJ (1989): The Yale Global Tic Severity Scale: initial testing of a clinician-rated scale of tic severity. *J Am Acad Child Adolesc Psychiatry* 28: 566–573.

2. Woods DW, Piacentini J, Himle MB, Chang S (2005): Premonitory Urge for Tics Scale (PUTS): initial psychometric results and examination of the premonitory urge phenomenon in youths with Tic disorders. *J Dev Behav Pediatr* 26: 397–403.

3. Sheehan DV, Sheehan KH, Shytle RD, Janavs J, Bannon Y, Rogers JE, *et al.* (2010): Reliability and validity of the Mini International Neuropsychiatric Interview for Children and Adolescents (MINI-KID). *J Clin Psychiatry* 71: 313–326.

4. Sheehan DV, Lecrubier Y, Sheehan KH, Amorim P, Janavs J, Weiller E, *et al.* (1998): The Mini-International Neuropsychiatric Interview (M.I.N.I.): the development and validation of a structured diagnostic psychiatric interview for DSM-IV and ICD-10. *J Clin Psychiatry* 59 Suppl 20: 22–33;quiz 34–57.

5. Wechsler D (n.d.): Wechsler Abbreviated Scale of intelligence. Retrieved July 20, 2023, from https://www.pearsonassessments.com/store/usassessments/en/Store/Professional-Assessments/Cognition-%26-Neuro/Wechsler-Abbreviated-Scale-of-Intelligence-%7C-Second-Edition/p/100000593.html

6. Busner J, Targum SD (2007): The Clinical Global Impressions Scale: Applying a Research Tool in Clinical Practice. *Psychiatry (Edgmont)* 4: 28.

7. Krishnan C, Santos L, Peterson MD, Ehinger M (2015): Safety of noninvasive brain stimulation in children and adolescents. *Brain Stimul* 8: 76–87.

8. Horowitz LM, Bridge JA, Teach SJ, Ballard E, Klima J, Rosenstein DL, *et al.* (2012): Ask Suicide-Screening Questions (ASQ): a brief instrument for the pediatric emergency department. *Arch Pediatr Adolesc Med* 166: 1170–1176.

9. Attkisson CC, Zwick R (1982): The client satisfaction questionnaire. Psychometric properties and correlations with service utilization and psychotherapy outcome. *Eval Program Plann* 5: 233–237.

10. DuBois M, Houlihan K, Raab B, Pryor A, Kellman M, Brinker M, *et al.* (2024): Quantifying tics: Best practices and design considerations for video-based tic coding in research. *Behav Res Methods* 56: 4073–4084.

11. Casey BJ, Cannonier T, Conley MI, Cohen AO, Barch DM, Heitzeg MM, *et al.* (2018): The Adolescent Brain Cognitive Development (ABCD) study: Imaging acquisition across 21 sites. *Dev Cogn Neurosci* 32: 43–54.

12. Drobyshevsky A, Baumann SB, Schneider W (2006): A rapid fMRI task battery for mapping of visual, motor, cognitive, and emotional function. *Neuroimage* 31: 732–744.

13. Wu SW, Maloney T, Gilbert DL, Dixon SG, Horn PS, Huddleston DA, *et al.* (2014): Functional MRI-navigated repetitive transcranial magnetic stimulation over supplementary motor area in chronic tic disorders. *Brain Stimul* 7: 212–218.

14. Nielsen AN, Greene DJ, Gratton C, Dosenbach NUF, Petersen SE, Schlaggar BL (2019): Evaluating the Prediction of Brain Maturity From Functional Connectivity After Motion Artifact Denoising. *Cereb Cortex* 29: 2455–2469.

15. Greene DJ, Koller JM, Hampton JM, Wesevich V, Van AN, Nguyen AL, *et al.* (2018): Behavioral interventions for reducing head motion during MRI scans in children. *Neuroimage* 171: 234–245.

16. Power JD, Mitra A, Laumann TO, Snyder AZ, Schlaggar BL, Petersen SE (2014): Methods to detect, characterize, and remove motion artifact in resting state fMRI. *Neuroimage* 84: 320–341.

17. Feczko E, Conan G, Marek S, Tervo-Clemmens B (2021): Adolescent Brain Cognitive Development (ABCD) Community MRI Collection and Utilities. *bioRxiv*. Retrieved from https://www.biorxiv.org/content/10.1101/2021.07.09.451638.abstract

18. Fair DA, Miranda-Dominguez O, Snyder AZ, Perrone A, Earl EA, Van AN, *et al.* (2020): Correction of respiratory artifacts in MRI head motion estimates. *Neuroimage* 208: 116400.

19. Glasser MF, Sotiropoulos SN, Wilson JA, Coalson TS, Fischl B, Andersson JL, *et al.* (2013): The minimal preprocessing pipelines for the Human Connectome Project. *Neuroimage* 80: 105–124.

20. Windhoff M, Opitz A, Thielscher A (2013): Electric field calculations in brain stimulation based on finite elements: an optimized processing pipeline for the generation and usage of accurate individual head models. *Hum Brain Mapp* 34: 923–935.

21. Opitz A, Paulus W, Will S, Antunes A, Thielscher A (2015): Determinants of the electric field during transcranial direct current stimulation. *Neuroimage* 109: 140–150.

22. Opitz A, Legon W, Rowlands A, Bickel WK, Paulus W, Tyler WJ (2013): Physiological observations validate finite element models for estimating subject-specific electric field distributions induced by transcranial magnetic stimulation of the human motor cortex. *Neuroimage* 81: 253–264.

23. Desikan RS, Ségonne F, Fischl B, Quinn BT, Dickerson BC, Blacker D, *et al.* (2006): An automated labeling system for subdividing the human cerebral cortex on MRI scans into gyral based regions of interest. *Neuroimage* 31: 968–980.

24. Seitzman BA, Gratton C, Marek S, Raut RV, Dosenbach NUF, Schlaggar BL, *et al.* (2020): A set of functionally-defined brain regions with improved representation of the subcortex and cerebellum. *Neuroimage* 206: 116290.

25. Piacentini J, Woods DW, Scahill L, Wilhelm S, Peterson AL, Chang S, *et al.* (2010): Behavior therapy for children with Tourette disorder: a randomized controlled trial. *JAMA* 303: 1929–1937.
